# Mathematical modelling of COVID-19 transmission dynamics with vaccination: A case study in Ethiopia

**DOI:** 10.1101/2022.03.22.22272758

**Authors:** Sileshi Sintayehu Sharbayta, Henok Desalegn Desta, Tadesse Abdi

**Author notes:** Correspondence should be addressed to Sileshi Sintayehu Sharbayta.

## Abstract

Mathematical modelling is important for better understanding of disease dynamics and developing strategies to manage rapidly spreading infectious diseases. In this work, we consider a mathematical model of COVID-19 transmission with double-dose vaccination strategy to control the disease. For the analytical analysis purpose we divided the model into two, model with vaccination and without vaccination. Analytical and numerical approach is employed to investigate the results. In the analytical study of the model we have shown the local and global stability of disease-free equilibrium, existence of the endemic equilibrium and its local stability, positivity of the solution, invariant region of the solution, transcritical bifurcation of equilibrium and sensitivity analysis of the model is conducted. From these analyses, for the full model (model with vaccination) we found that the disease-free equilibrium is globally asymptotically stable for *R*_*v*_ < 1 and is unstable for *R*_*v*_ > 1. A locally stable endemic equilibrium exists for *R*_*v*_ > 1, which shows the persistence of the disease if the reproduction parameter is greater than unity. The model is fitted to cumulative daily infected cases and vaccinated individuals data of Ethiopia from May 01, 2021 to January 31, 2022. The unknown parameters are estimated using the least square method with the MATLAB built-in function ‘lsqcurvefit’. The basic reproduction number, *R*_0_ and controlled reproduction number *R*_*v*_ are calculated to be *R*_0_ = 1.17 and *R*_*v*_ = 1.15 respectively. Finally, we performed different simulations using MATLAB. From the simulation results, we found that it is important to reduce the transmission rate, infectivity factor of asymptomatic cases and, increase the vaccination coverage and quarantine rate to control the disease transmission.

## 1. Introduction

Coronavirus (COVID-19) is an infectious disease caused by a novel coronavirus, which is a respiratory illness that can spread in a population in several different ways. A person can be infected when droplets containing the virus are inhaled or come directly into contact with the eyes, nose or mouth. The novel coronavirus has been spreading worldwide starting from the first identification in December 2019. The world health organization (WHO) declared COVID-19 as pandemic on March 12, 2020. From the first day of the outbreak to February 21, 2023, more than 757.2 million confirmed cases and more than 6.8 million confirmed deaths are registered worldwide [32]. The same report shows 499833 confirmed cases and 7, 572 confirmed deaths in the same period of time in Ethiopia.

The world is struggling to control the pandemic by imposing different restrictions based on country-specific strategies. Besides the restrictions, nowadays different countries are delivering vaccines to their people. As of 21 February 2023, 11 vaccines were granted for emergency use by WHO [31]. These are Novavax, COVOVAX, Moderna, Pfizer/BioNTech, Janssen (Johnson & Johnson), AstraZeneca, Vaxzevria (Oxford/AstraZeneca), Covishield (Oxford/AstraZeneca formulation), Covaxin, Sinopharm and Sinovac. Country approvals of this vaccine varies. For example, Pfizer/BioNTech and Oxford/AstraZeneca are approved by 149 countries, Janssen (Johnson & Johnson) is approved by 113, and Moderna is approved by 88 countries worldwide [31]. Until February 18, 2023, about 13.2 billion COVID-19 vaccine doses are administered globally. The portion 69.6% of the world population have received at least one dose of COVID-19 vaccine and this coverage represents developed counties due to scarcity of the vaccine in low-income countries. Only 27.6% of people in low-income countries have received at least one dose [26]. Up to 21 January 2023, a total of 53, 514, 115 vaccine doses have been administered in Ethiopia [32].

Studies involving mathematical models of infectious disease are helping the public health authorities by giving them an in-depth information through analysis of dynamics of the disease to make an informed decisions and policy making. Oftentimes, deterministic models based on classical derivatives are used to study the disease transmission dynamics. These studies are also powerful tools for predicting the future aspects of a disease. As far as COVID-19 is concerned, currently there are several such researches which have been conducted and are helping the struggle towards containing the spread.

Before vaccines are produced, mathematical models for COVID-19 focused on assessing the impacts of non-pharmaceutical interventions(NPIs) like social distancing, wearing masks, personal hygiene, partial or full lockdown and the like as control strategies. For the details on this we mention [25, 1, 29, 3, 22, 13, 4] and the references therein. Mathematical model of SARS-CoV-2 transmission with optimal control is studied in [25] using the data from USA and they found that a major factor that differentiates strategies that prioritize lives saved versus reduced time under control is how quickly control is relaxed once social distancing restrictions expire. They also highlighted that the scope of controlling the COVID-19 until vaccines are available depends on epidemiological parameters. The study in [29], which studies the transmission of COVID-19 in crowded settlements revealed that level of compliance to standard operating procedures (SOPs) (such as use of masks, physical distancing measures and effective contact tracing) increases, then the disease prevalence peaks are greatly reduced and delayed. Authors in [3] studied a model of the transmission dynamics of corona virus disease in India focusing on basic non-pharmaceutical interventions. Their results showed that the implementation of an almost perfect isolation in India and 33.33% increment in contact-tracing on June 26, 2020 may reduce the number of cumulative confirmed cases of COVID-19 by around 53.8% at the end of July 2020. In [1], modifiying the Kermack–McKendrick SEIR model the authors studied the population-level impact of implementing behavioural change control measures, the time horizon necessary to reduce the effective contact rate, and the proportion of people under sanitary emergency measures in controlling COVID-19 in Mexico. Simulation results of this paper indicated that the most likely dates for maximum incidence happen under a scenario of high Sanitary Emergency Measures (SEM) compliance and low SEM abandonment rate. Even if the quality of the face mask is frequently questioned, wearing a face mask is one of the non-pharmaceutical measures. The study in [13] suggests that broad adaption of even the relatively ineffective face masks may significantly reduce the transmission and hospitalization peak and death. For combating COVID-19, the timing of relaxing or lifting of non-pharmaceutical measures is essential. From this point of view, the authors in [22] showed the crucial importance of relaxation or lifting of strict social distancing measures in determining the future aspect of COVID-19 pandemic.In particular one of their results show shows that early termination of the strict social-distancing measures could trigger a devastating second wave with burden similar to those projected before the onset of the strict social-distancing measures were implemented. In [4], they evaluate and compare the effectiveness of the four types of NPIs of COVID-19, namely: the implementation of a mandatory mask, quarantine or isolation, distancing and traffic restriction in 190 countries between 23 January and 13 April 2020. In their study, they indicated that NPIs could significantly hold the COVID-19 pandemic. Social distancing and the implementation of two or more NPIs should be the priority strategies for holding COVID-19.

Forecasting the COVID-19 pandemic is crucial for health care planning and controlling the disease. In this respect, the authors in [16] proposed a COVID-19 model with contact tracing and hospitalization strategies and performed short-term and long-term predictions for daily and cumulative confirmed cases of COVID-19 outbreaks for five provinces of India. In the short-term predictions, some states show exponential growth and others show decay of daily new cases. Long term predictions for India show to exhibit oscillatory dynamics. A COVID-19 model in [27] predicts the dynamics of COVID-19 in 17 provinces of India and overall India. One of the results in this study shows that combining the restrictive social distancing and contact tracing will make the elimination of COVID-19 pandemic possible.

Currently, vaccines are available as one of the main control strategies. Epidemiological modelers started to incorporate this additional intervention to see the dynamic properties of the disease and sort out some important policy directions for the public health authorities. In this regard, there are a number of studies, from which [10, 21, 6, 15] can be mentioned. A mathematical model of COVID-19 with comorbidity was formulated to study the transmission dynamics and an optimal control-based framework to mitigate the disease transmission in [10]. In this study, the authors found that disease persists with the increase in exposed individuals having comorbidity in society and an optimal strategy with combined measures provides effective protection of the population with minimum social and economic costs. Even during vaccination, non-pharmaceutical interventions are essential: In this regard the study in [21] showed that relaxing restrictions would cause benefits from vaccination to be lost by increasing case numbers and hence vaccination alone is insufficient to contain the outbreak. Another problem in attaining herd immunity in the population is vaccine hesitancy in the event that vaccination is not mandatory, in which case people are the last to decide either to get vaccinated or otherwise. A behavioural modelling approach was used to assess the impact of hesitancy and refusal of vaccine on the dynamics of the COVID-19 [6]. In this paper, the authors showed hesitancy and refusal of vaccination is better contained in case of large information coverage and small memory characteristics. In the study [15] the author analyzed the onset of COVID-19 spread in countries such as China, Italy, Spain, the United States, the United Kingdom, Japan, France, and Germany based on publicly available statistical data aiming to establish the laws of the spread of COVID-19 and to use them to develop a mathematical model to predict changes and make informed control policy decisions. In the study specific values for SARS-CoV-2 transmissibility and COVID-19 duration were estimated for different countries. It was found that in China, the viral transmissibility was3.12 before quarantine measures were implemented and 0.36 after these measures were lifted. For the other countries, the viral transmissibility was 2.28 − 2.76 initially, and it then decreased to 0.87 − 1.29 as a result of quarantine measures. Therefore, it can be expected that the spread of SARS-CoV-2 will be suppressed if 56% − 64% of the total population becomes vaccinated or survives COVID-19.

Even with these immunizations, the virus continues to spread in many countries, with some vaccinated people becoming infected, necessitating the delivery of booster shots. Recently, authors in [24, 2] have built mathematical models devoted to studying the impact of double dosage vaccination. The authors of [24] looked at a COVID-19 model with a double-dose vaccination strategy to reduce the illness outbreak in Bangladish. According to the findings, a full-dose vaccination campaign has the ability to eradicate the virus from the community. A similar study [2] was undertaken for the case in Ghana, and it revealed that implementing double-dose vaccination and quarantine will help reduce the spread of COVID-19. We will consider a similar model with double-dose vaccination in the the case of Ethiopia.

A few epidemiological modeling studies of COVID-19 based on Ethiopian data have been undertaken, and we will highlight some of them here. In [20], the authors considered a mathematical model for the transmission dynamics of COVID-19 by incorporating self-protection behavioral changes in the population. Based on the available data from Ethiopia and other countries, they estimated the unknown parameter values using a combination of least squares and Bayesian estimation methods. They found that the sensitive parameters for the spread of the virus vary from country to country and control of the effective transmission rate (recommended human behavioral change towards self-protective measures) is essential to stop the spread of the virus. A mathematical model of COVID-19 in the case of Ethiopia is also considered in [17].Indeed in this study they found that the spread of COVID-19 can be managed by minimizing the contact rate of infected and increasing the quarantine of exposed individuals. There is also another COVID-19 mathematical modelling for optimal control and assessing the impact of nonpharmaceutical interventions on the dynamics of COVID-19 which are specific to Ethiopian data [11, 14]. Even with vaccines in place as an intervention for the COVID-19 pandemic, countries are still struggling to control the disease. Better understanding of disease dynamics and forecasting will be paramount for developing better pandemic management strategies. We also believe that scientific studies on COVID-19 transmission in Ethiopia are limited and that, as far as we reviewed, no mathematical modeling studies have been conducted in light of the current situation (including double-dose vaccination). As a result, we consider a mathematical model of COVID-19 transmission dynamics with double-dose vaccination in our study.

The paper is organized as follows: In Section (2), we describe the model and formulate the pertinent differential equation. In Section (3), we carry out mathematical analysis of the model. Section (4) is devoted to numerical simulation and discussion. In Section (5), we present prediction of the cumulative vaccine administered with respect to the first dose vaccination rate. Finally, in Section (6) the conclusion is presented.

## 2. Model description and formulation

In this study, we proposed a model where the total population is divided into nine compartments. Namely Susceptible, are uninfected people with the disease but have a chance to be infected; Vaccinated with first dose, but still have the chance to be infected; vaccinated with second dose, individuals who completed the two doses within the specified time; Exposed, Infected but not yet infectious; Asymptomatic infectious, people who are infected but does not show symptoms but have the chance to transmit the disease; Symptomatic infectious, are those who are infected and show symptoms; Quarantine, are individuals who are tested positive so that isolated from the population; Hospitalized, are those who are in critical health and joined hospitals for treatment; and Recovered, Recovered from the disease; denoted by *S, V*_1_, *V*_2_, *E, I*_*a*_, *I*_*s*_, *Q, H* and *R* respectively. We assumed that individuals in *Q* and *H* compartments are isolated from the population and hence they will have negligible role in transmitting the disease. Therefore, only individuals in *I*_*a*_ and *I*_*s*_ are capable of transmitting the disease. Vaccines available for COVID-19 do not totally prevent infection. Thus individuals in *S, V*_1_ and *V*_2_ compartments can get infected with the force of infection 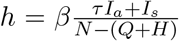. Such a force of infection is used in most COVID-19 models [17, 11, 5], where *β* is the transmission rate, *τ* is the infectivity factor of asymptomatic individuals and *N* is the total population. Due to the vaccine, individuals in *V*_1_ and *V*_2_ classes are relatively less infected than the fully susceptible ones and they will get infected with reduced vulnerability of (1 − *η*_1_) and (1 − *η*_2_) respectively. The quantities *η*_1_ and *η*_2_ measure the effectiveness of the first dose and the second dose vaccine respectively. Majority of the vaccines approved by WHO are given in two doses with an average recommended time interval between the two doses. We considered this scenario in our model. Susceptible individuals get vaccination (the first dose) at the rate of *p*_1_ and those who got the first dose will get the second dose after an average 1*/α* period of time with the rate of *p*_2_. In this study we did not fix a particular vaccine type therefore the value of 1*/α* represents the average time needed to take the second dose. From the population, *ρ* proportion of exposed individuals will move to the asymptomatic class and the rest, (1 − *ρ*) proportion will move to the symptomatic class after they finish the incubation period of 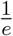 day, where *e* is the infection rate. Mostly the symptoms of COVID-19 are similar to other respiratory diseases like common cold and flue, so all symptomatic individuals are not quarantined. Those only tested and confirmed can go to quarantine. Symptomatic individuals get tested and quarantined at the rate of *δ*. Those quarantined may develop serious illness, in this case they go to hospital at the rate of *q*_*h*_. Individuals in *I*_*a*_, *I*_*s*_, *Q* and *H* will recover from the disease at the rate of *r*_*a*_, *r*_*s*_, *r*_*q*_ and *r*_*h*_ respectively and they are presumed to be immunized for the rest of their lives once they have recovered. Asymptomatic individuals are with less pain and assumed does not show symptoms and will not die due to the disease. As a consequence, individuals in *I*_*s*_, *Q* and *H* classes die due to the disease at the rate of *d* (assumed to be equal). People in all compartments will die naturally at the rate of *μ* and the quantity *π* is the recruitment rate to the susceptible compartment. The total population size at time *t* is denoted by *N*(*t*) where,

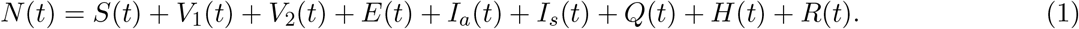

The model flow diagram is shown in Figure 1.

**Figure 1:**
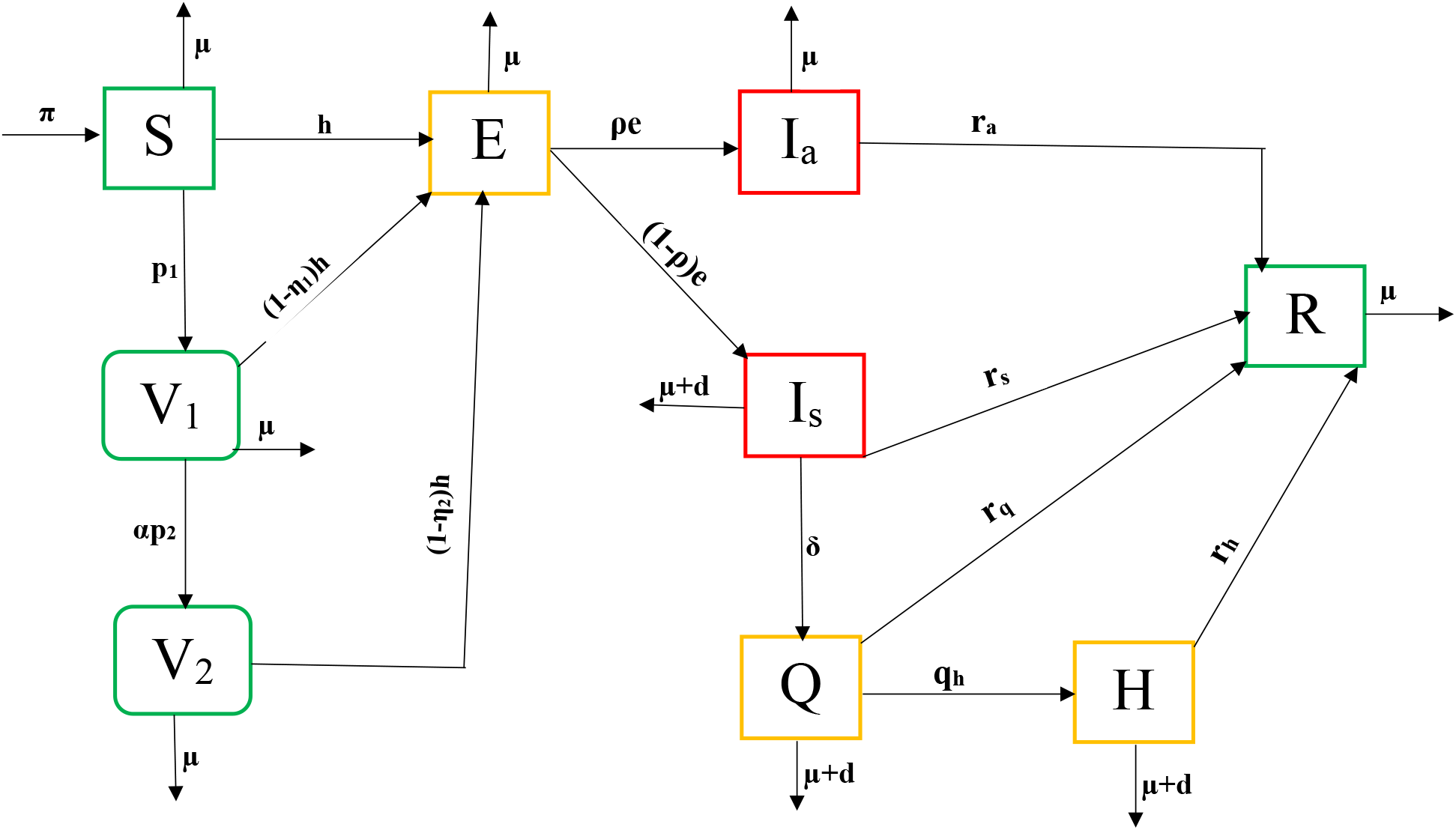
Disease transmission diagram: green compartment indicates non-infected, the red compartment is infected and infectious and the yellow compartment shows infected but assumed to be not infectious(*Q* and *H*), on incubation period (*H*).

From the schematic diagram Figure(1) the following system of differential equations is obtained

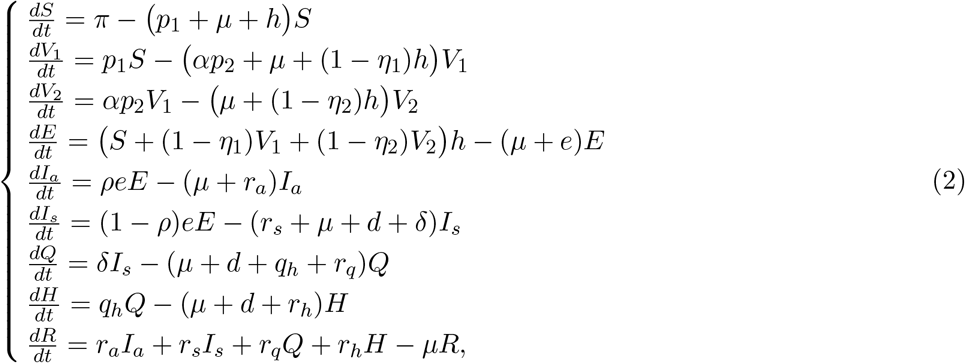

with initial conditions

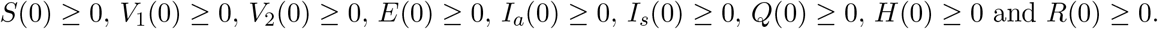

## 3. Model analysis

In this section, positivity of solution, the invariant region, production number, stability analysis of disease-free and endemic equilibrium point, bifurcation and sensitivity analysis are discussed.

### 3.1 Positivity and boundedness of the solutions

Since each component of the given model system considers a human population, it is necessary to show that all variables *S*(*t*), *V*_1_(*t*), *V*_2_(*t*), *E*(*t*), *I*_*a*_(*t*), *I*_*s*_(*t*), *Q*(*t*), *H*(*t*) and *R*(*t*) are positive for all *t* > 0.

#### Theorem 3.1.1

*If S*(0) ≥ 0, *V*_1_(0) ≥ 0, *V*_2_(0) ≥ 0, *E*(0) ≥ 0, *I*_*a*_(0) ≥ 0, *I*_*s*_(0) ≥ 0, *Q*(0) ≥ 0, *H*(0) ≥ 0 *and R*(0) ≥ 0, *then the solution set* {*S*(*t*), *V*_1_(*t*), *V*_2_(*t*), *E*(*t*), *I*_*a*_(*t*), *I*_*s*_(*t*), *Q*(*t*), *H*(*t*), *R*(*t*)} *of the model* (2) *consists of positive members for all t* > 0.

*Proof*. From the first equation of the system (2), we have

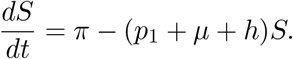

This leads to,

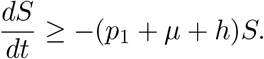

And hence,

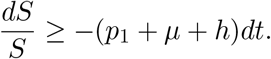

Finally upon integration, we obtain,

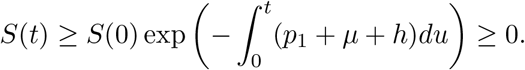

Thus, *S*(*t*) ≥ 0.

Similarly, it can be shown that the other equations of system (2) are positive for all *t* > 0. Hence, the state variables of the system are all positive for all *t* > 0.

#### Theorem 3.1.2

*The feasible solution set* {*S, V*_1_, *V*_2_, *E, I*_*a*_, *I*_*s*_, *Q, H, R*} *of the model* (2) *with the given initial condition remains bounded in the region* 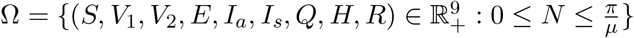.

*Proof*. Differentiating *N* in equation (1) with respect to *t* we obtain;

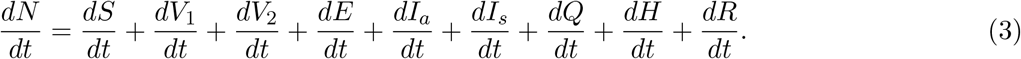

Using system (2) and evaluating at (3) gives us;

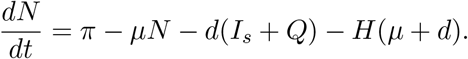

Since the state variables of system *I*_*s*_, *Q* and *H* are positive for all *t* ≥ 0 we have

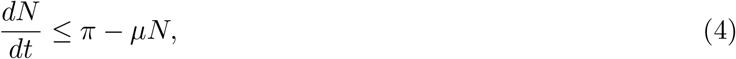

in which *N* is asymptotically bounded

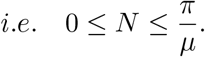

This completes the proof. □

### 3.2 Reproduction number, existence and stability analysis of equilibria

#### 3.2.1 Disease-free equilibrium point

In this subsection, we determine the equilibrium point at which there is no disease in the population (i.e. *I*_*a*_ = *I*_*s*_ = *Q* = *H* = *E* = *R* = 0) by setting the right hand side of system (2) to zero. We get:

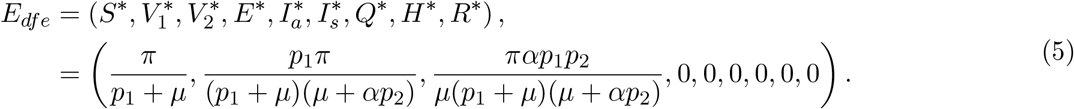

##### Remark 1

*In* (5), *when there is no vaccination, i*.*e*., *p*_1_ = 0, *the disease-free equilibrium will be reduced to a fully susceptible disease-free state given by*

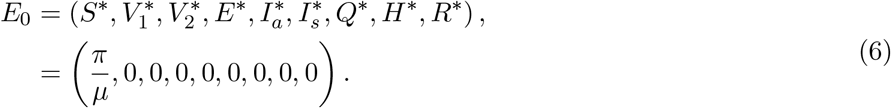

*If p*_1_ = 1 *we get a disease-free equilibrium in which every susceptible individual is vaccinated with the first dose, which can be expressed as*

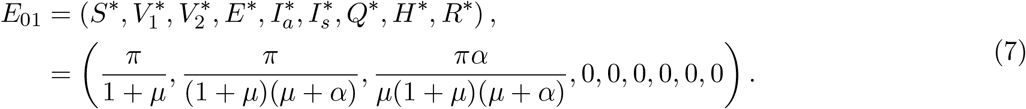

#### 3.2.2 Reproduction number

The basic reproduction number (*R*_0_) is the average number of secondary cases produced by one primary infection during the infectious period in a fully susceptible population and the control reproduction number (in our case denoted by *R*_*v*_) is used to represent the same quantity for a system incorporating control (or intervention) strategies [30]. We will use the next generation matrix method [12] to find the basic and control reproduction number.

Let the matrix for new infection appearance at the infected compartment be given by ℱ,

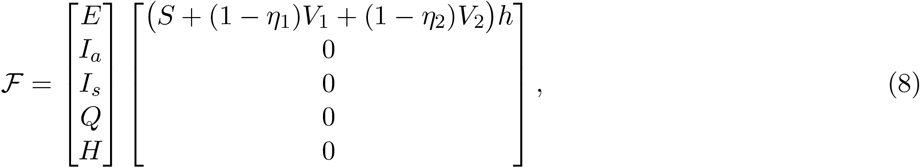

and the matrix of other transactions at each of the infected compartments can be represented by 𝒱, and is given by

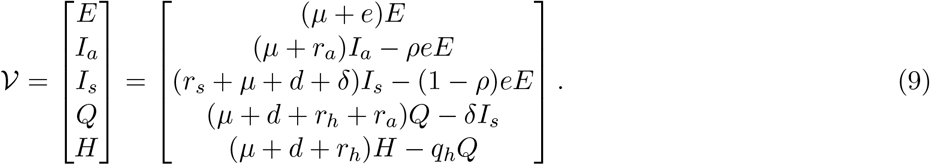

Now finding the Jacobian of ℱ and 𝒱, we get the matrices *F* (only the first row, nonzero row) and *V* written as;

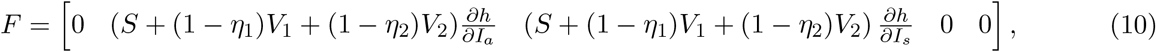

where,

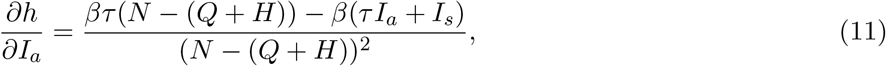

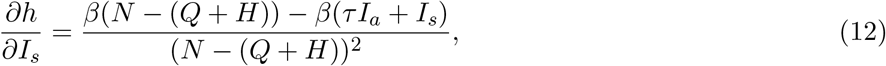

and

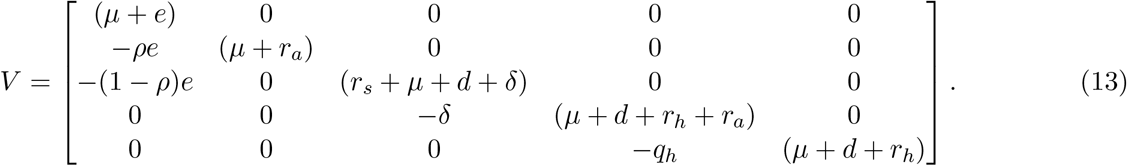

The control reproduction number is given by *R*_*v*_ = *ν*(*F*(*E*_*dfe*_) × *V* ^−1^). Where *ν* is the spectral radius of the matrix *F*(*E*_*dfe*_) × *V* ^−1^. Thus *R*_*v*_, can be written as:

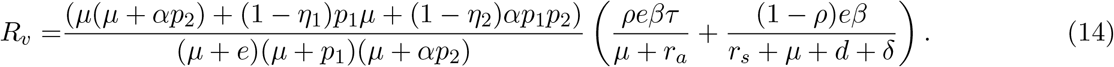

The basic reproduction number, *R*_0_ is obtained by setting *p*_1_ = *p*_2_ = 0 in (14) and is given by:

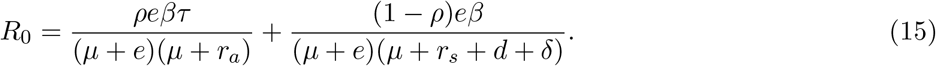

We can rewrite equation (14) in terms of *R*_0_ as;

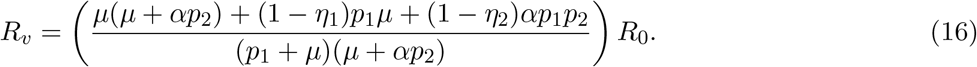

In system (2), the solution for the state variables *Q, H* and *R* can easily be found from other variables in the system and they do not affect them. Therefore in the following subsections we restrict our mathematical analysis to the following system of equations.

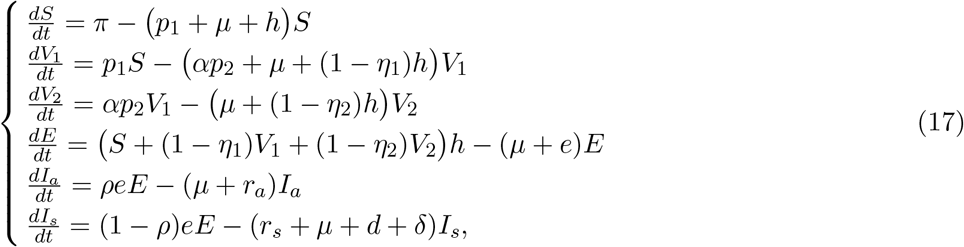

### 3.3 Model without vaccination

In this subsection we will study the reduced model system (17) when there is no vaccination (*p*_1_ = 0 = *p*_2_). which will further be reduced to a system represented in the following equation,

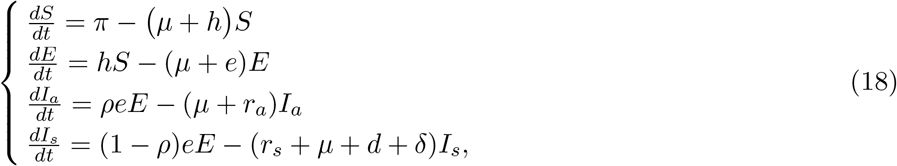

For the model (18) the reproduction number can be found by replacing *p*_1_ = 0 = *p*_2_, which is the basic reproduction number of the full model, and it is as given equation (15) and the disease free equilibrium is as written in (6). Consequently, we have the following result

#### Theorem 3.3.1

*The disease free equilibrium E*_0_ *is locally asymptotically stable if R*_0_ < 1 *and unstable if R*_0_ > 1.

*Proof*. The Jacobian matrix of the system (18) is given by:

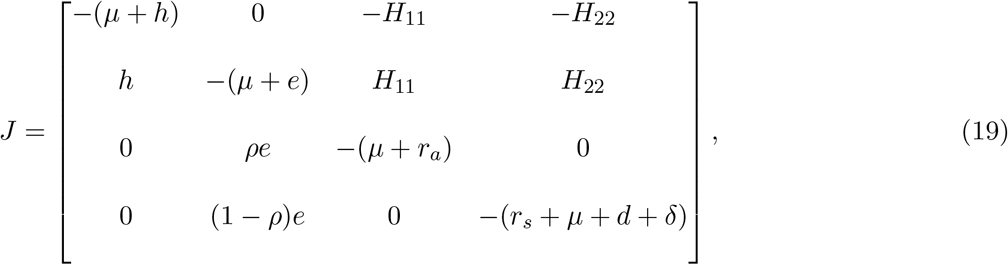

where

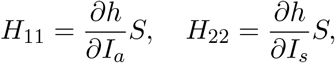

and 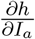 and 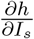 are as in equations (11) and (12) respectively.

The Jacobian matrix (19) evaluated at the disease-free equilibrium (*E*_0_) is given by:

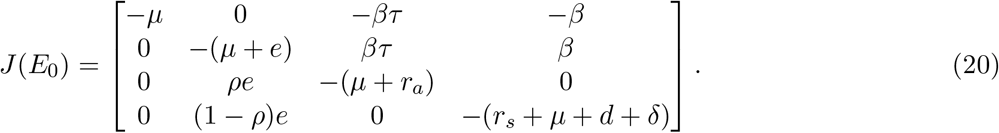

The characteristic equation of the matrix (20) is given by

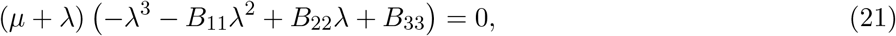

where

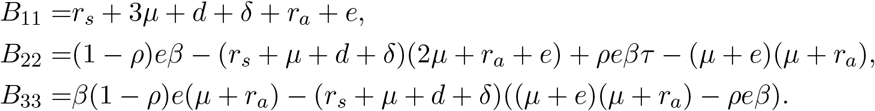

From (21) we have the roots given by *λ*_1_ = −*μ* and −*λ*^3^ − *B*_11_*λ*^2^ + *B*_22_*λ* + *B*_33_ = 0. By Descartes’ rule of sign, the roots of the later equation will be negative if *B*_22_ < 0 and *B*_33_ < 0.

Suppose *R*_0_ < 1. This implies that

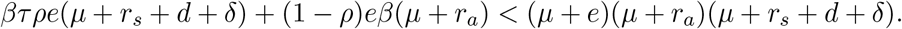

Therefore,

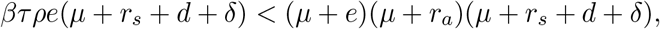

and

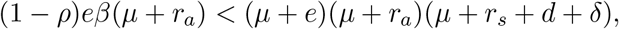

which are equivalently written as

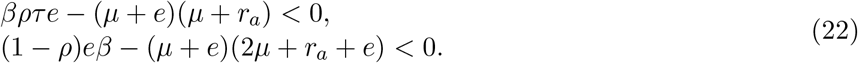

From the inequalities in (22), it can be concluded that *B*_22_ < 0 if *R*_0_ < 1. Similarly it can be shown that *B*_3_ < 0 whenever *R*_0_ < 1. Therefore, the disease-free equilibrium *E*_0_ is locally asymptotically stable if *R*_0_ < 1. For *R*_0_ > 1, *B*_22_ will be positive. And hence we will have at least one positive eigenvalue. Thus, *E*_0_ will be locally unstable. □

#### 3.3.1 Global stability of disease-free equilibrium

For we seek to investigate the global stability of disease-free equilibrium, we use the technique implemented by Castillo-Chavez et al. [8]. To implement the technique we write our model system in the form:

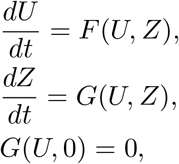

where *U* represents an uninfected compartment and *Z* represents infected compartment. Thus, the disease-free equilibrium point of the model can be represented by *U*^*^ = (*U*_0_, 0). Thus, for *R*_0_ < 1, for which the disease-free equilibrium point is locally asymptotically stable the following two conditions are sufficient to guarantee the global stability of disease-free equilibrium point (*U*_0_, 0).

(H1) For 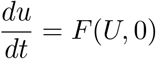, *U*_0_ is globally asymptotically stable.

(H2) 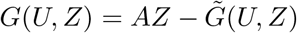, where 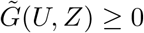 for all (*U, Z*) ∈ Ω where *A* = *D*_*I*_*G*(*U*_0_, 0) is a M-matrix (the off-diagonal elements of *A* are nonnegative) and Ω is the region where the model makes biological sense.

##### Theorem 3.3.2

*The disease-free equilibrium point E*_0_ *is globally asymptotically stable provided that R*_0_ < 1.

*Proof*. For the system (18) we have 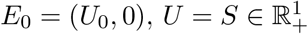 and 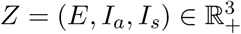. For condition (H1), *F*(*U, Z*) can be written as

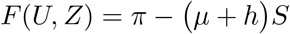

Hence,

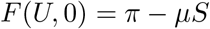

It is obvious that 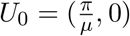 is globally asymptotically stable for *F*(*U*, 0).

For condition (H2), from the system (18) we can get *G*(*U, Z*),

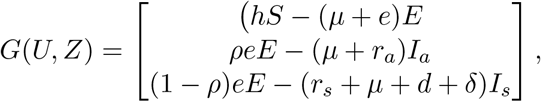

and the M-matrix is calculated as

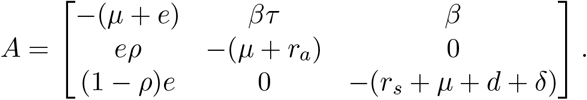

Thus we have,

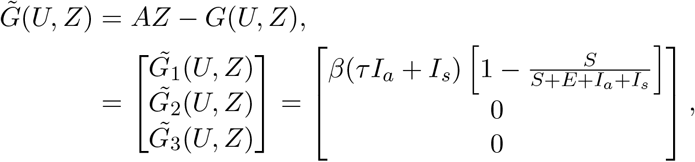

which leads to 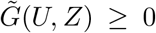 for all (*U, Z*) ∈ Ω. Hence both the conditions (H1) and (H2) are satisfied.

Therefore, by Castillo-Chavez et al. [8] the disease-free equilibrium point is globally asymptotically stable for *R*_0_ < 1. □

The endemic equilibrium of the model with no vaccination (18) is calculated as

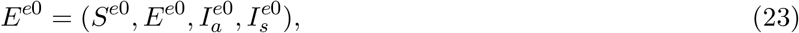

and the components are given by,

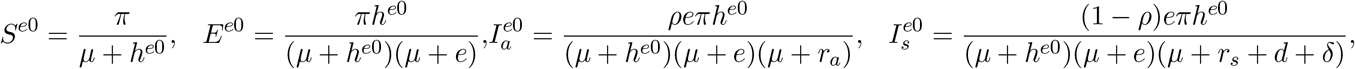

where

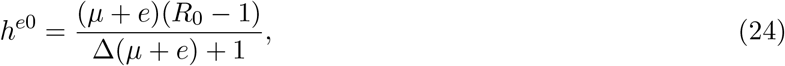

and

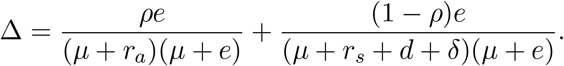

From equation (24) *h*^*e*0^ > 0 if and only if *R*_0_ > 1, therefore we have the following result.

##### Lemma 3.3.1

*The system* (18) *have a unique endemic equilibrium if R*_0_ > 1 *and have no endemic equilibrium for R*_0_ < 1.

The characteristic equation of the Jacobian matrix (19) evaluated at the endemic equilibrium (24) is obtained as

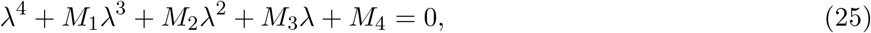

where

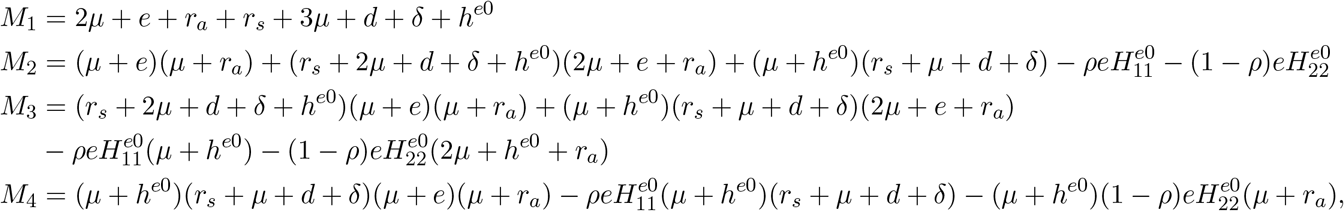

and 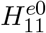 and 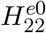 are values of *H*_11_ and *H*_22_ evaluated at the endemic equilibrium respectively. Since *M*_1_ > 0 by Descartes rule of sign the characteristic equation (25) will have negative roots if *M*_2_ > 0, *M*_3_ > 0 and *M*_4_ > 0. Therefore we have the following result.

##### Theorem 3.3.3

*The endemic equilibrium* (23) *is locally asymptotically stable if R*_0_ > 1. *and M*_2_ > 0, *M*_3_ > 0 *and M*_4_ > 0.

### 3.4 Model with Vaccination

In this subsection will consider the system with vaccination (17) and present its mathematical analysis.

#### 3.4.1 Local stability of disease-free equilibrium

##### Theorem 3.4.1

*The disease-free equilibrium, E*_*dfe*_ *is locally asymptotically stable if R*_*v*_ < 1 *and unstable if R*_*v*_ > 1.

*Proof*. The Jacobian matrix of the system (17) is given by:

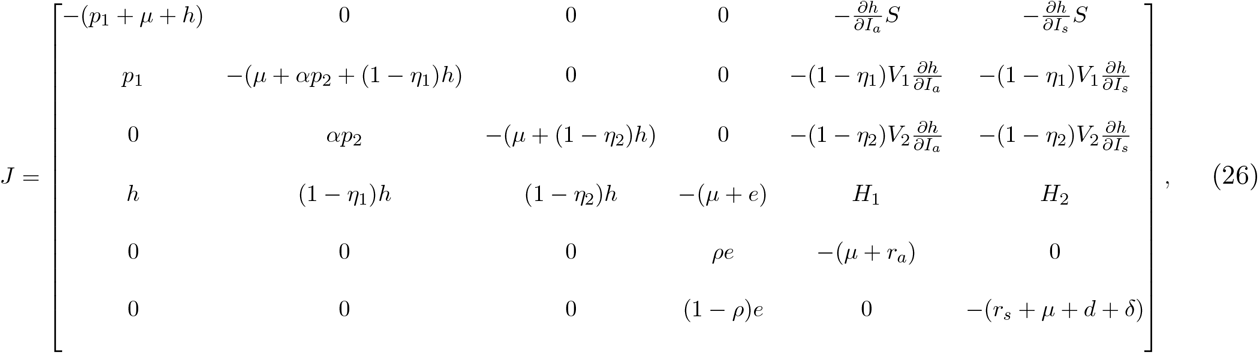

where

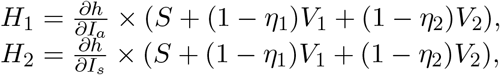

and 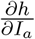 and 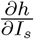 are as in equations (11) and (12).

The Jacobian matrix (26) evaluated at the disease-free equilibrium *E*_*dfe*_ is given by:

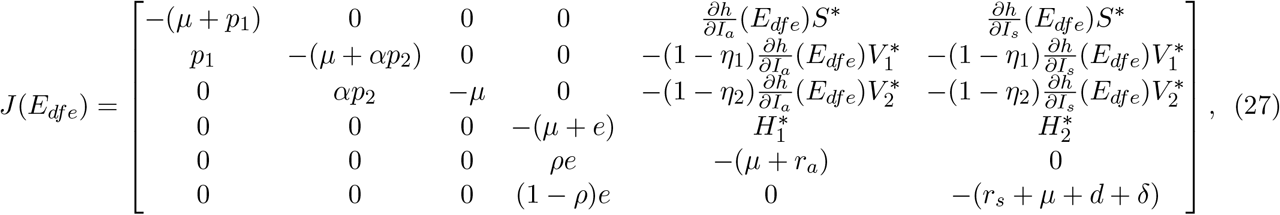

where

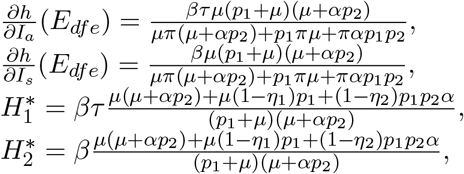

and its characteristic equation is:

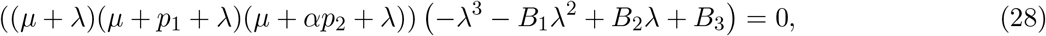

where

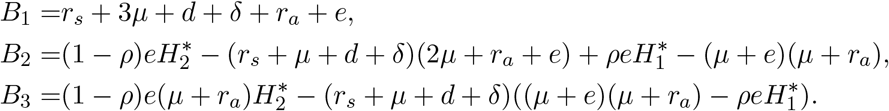

From (28) we have the roots given by *λ*_1_ = −*μ, λ*_2_ = −(*μ* + *αp*_2_), *λ*_3_ = −(*μ* + *p*_1_) and −*λ*^3^ − *B*_1_*λ*^2^ + *B*_2_*λ* + *B*_3_ = 0. By Descartes’ rule of sign, the roots of the later equation will be negative if *B*_2_ < 0 and *B*_3_ < 0.

Let us write the equation for *R*_*v*_ in (14) in terms of 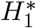 and 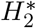 as:

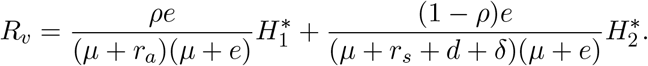

Suppose *R*_*v*_ < 1. This implies that

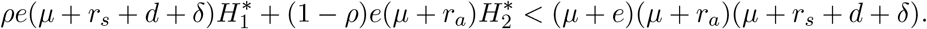

Therefore,

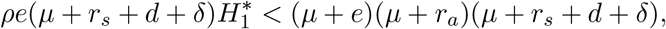

and

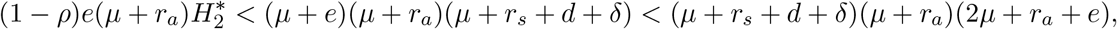

which are equivalently written as

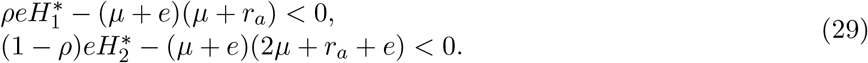

From the inequalities in (29), we summarize that *B*_2_ < 0 if *R*_*v*_ < 1. And it can also be shown that *B*_3_ < 0 whenever *R*_*v*_ < 1. Therefore, the disease-free equilibrium *E*_*dfe*_ is locally asymptotically stable if *R*_*v*_ < 1. For *R*_*v*_ > 1, *B*_2_ will be greater than zero. And hence we will have at least one positive eigenvalue. Thus, *E*_*dfe*_ will be unstable.

#### 3.4.2 Global stability of disease-free equilibrium point

We use the method implemented in section 3.3.1 to show the global stability. Let 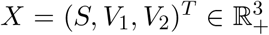 be represent uninfected individual and 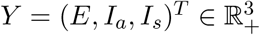 be represent infected compartments.

##### Theorem 3.4.2

*The point E*_*dfe*_ = (*X*\*, 0) *is globally asymptotically stable provided that R*_*v*_ < 1 *and* 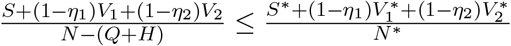.

*Proof*. For condition (H1) from the system (17) we can get *F*(*X, Y*), i.e.

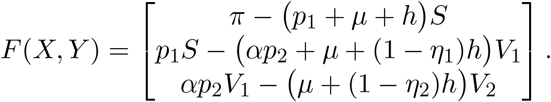

Hence,

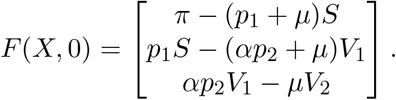

It is obvious that 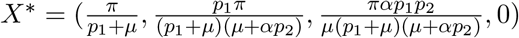 is globally asymptotically stable for *F*(*X*, 0) as *X* → *X*\* when *t* → ∞.

For condition (H2), from the system (17) we can get *G*(*X, Y*),

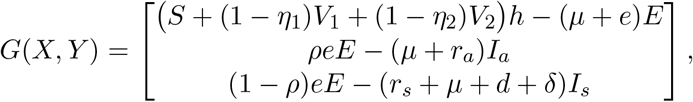

and

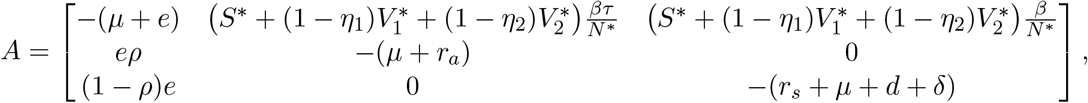

where,

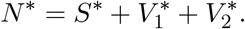

We have,

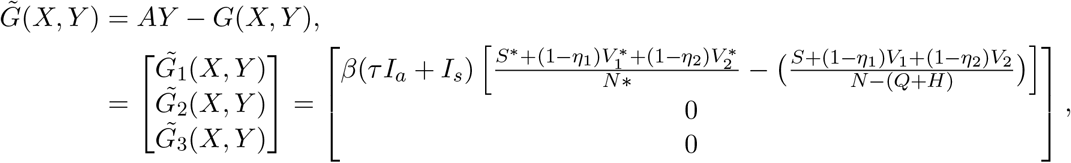

thus,

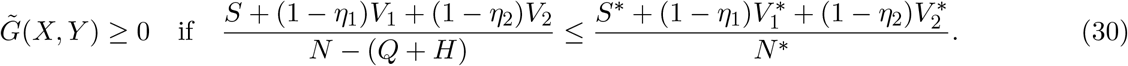

Therefore, the disease-free equilibrium point is globally asymptotically stable for *R*_*v*_ < 1 and the condition given in equation 30 is satisfied. □

#### 3.4.3 Existence of endemic equilibrium

By equating the system (2) to zero, we get the endemic equilibrium in terms of the force of infection *h* and we denote it by,

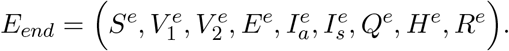

The components of *E*_*end*_ are given as follows:

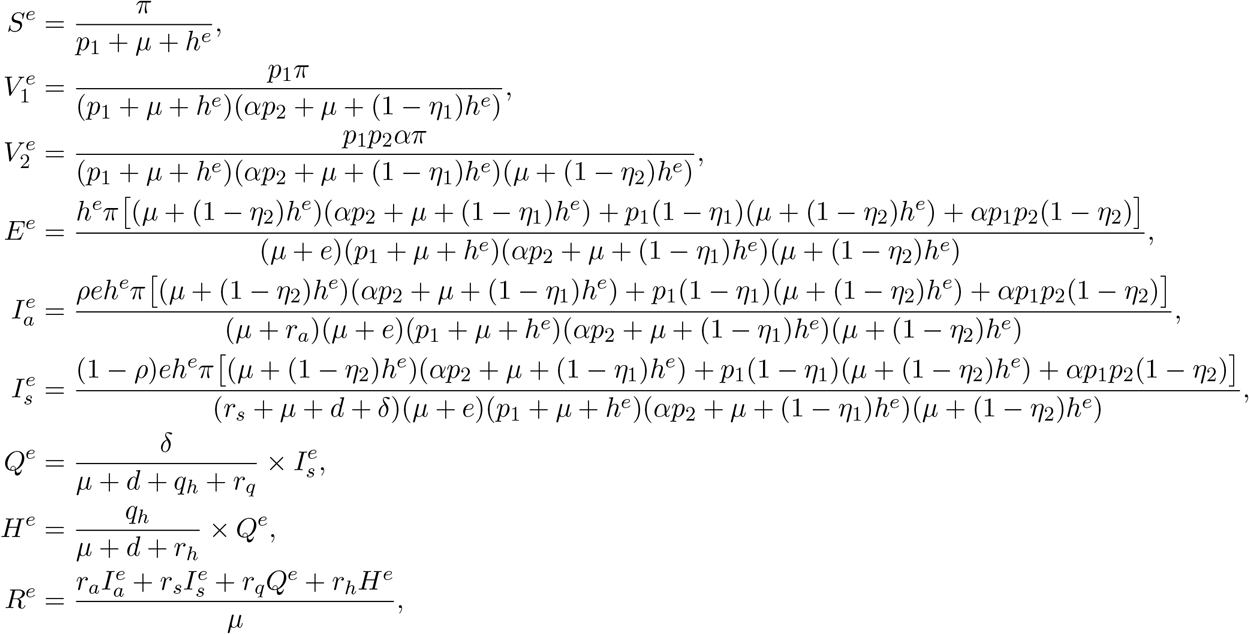

where *h*^*e*^ is the positive root of the equation

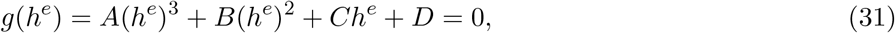

obtained from

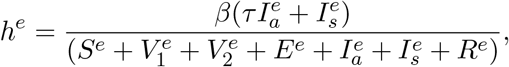

and the coefficients in equation (31) are given by:

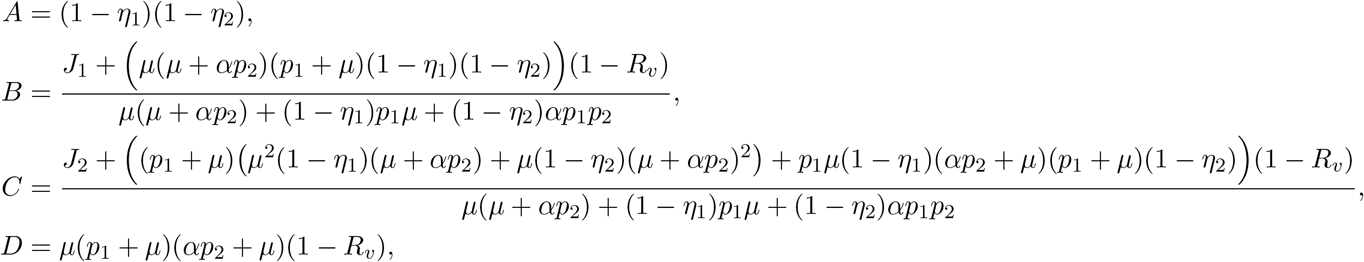

where,

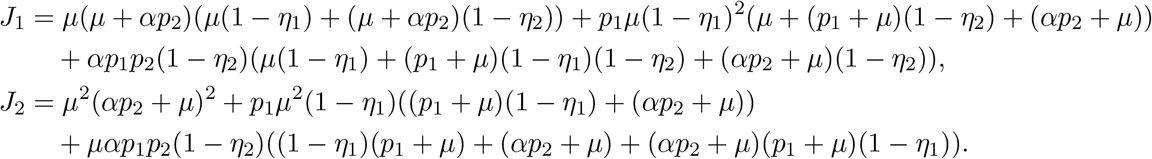

It can easily be seen that *A* > 0. If *R*_*v*_ > 1 then *D* < 0, therefore *h*(0) < 0. Additionally lim 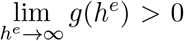. Therefore, from the continuity of *g*, there exists at least one positive 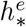 such that 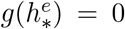 and hence there will be at least one endemic equilibrium of the model system (2). On the other hand, if *R*_*v*_ < 1, then *B* > 0, *C* > 0 and *D* > 0 then by Descartes’ rule of sign, (31) has no positive real root, which proves that there is no endemic equilibrium point when *R*_*v*_ < 1. From the above discussion, we can state the following theorem.

##### Theorem 3.4.3

*If R*_*v*_ > 1, *there exists at least one endemic equilibrium point for the model system* (2) *and there is no endemic equilibrium point for the model system* (2) *when R*_*v*_ < 1.

### 3.5 Bifurcation analysis

We will use the approach in [7] to determine the occurrence of a trasncritical bifurcation at *R*_*v*_ = 1. The method relies on the general center manifold theory, where the normal form representing the dynamics of the system on the central manifold is given by:

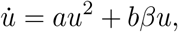

with

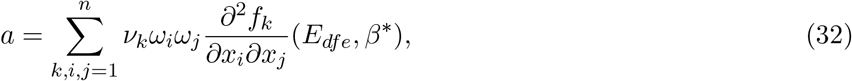

and

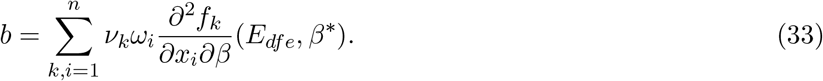

Here *β* has been chosen as a bifurcation parameter and *β*\* is its critical value, *f* represents the right–hand side of the system (17), *x* represents the state variable vector, *x* = (*x*_1_, *x*_2_, *x*_3_, *x*_4_, *x*_5_, *x*_6_) = (*S, V*_1_, *V*_2_, *E, I*_*a*_, *I*_*s*_), *ν* and *ω* are the left and right eigenvectors corresponding to the zero eigenvalue of the Jacobian matrix at the disease-free equilibrium and the critical value, i.e., at *E*_*dfe*_ and *β* = *β*\*. When *R*_*v*_ = 1, which is equivalent to *β* = *β*\*, with

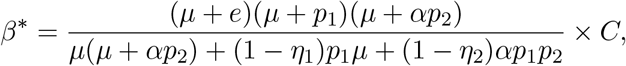

where,

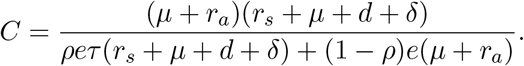

Thus, according to Theorem 4.1[7], the disease-free equilibrium is locally asymptotically stable if *β* < *β*\*, and it is unstable when *β* > *β*\*. The direction of the bifurcation occurring at *β* = *β*\* can be derived from the sign of the coefficients (32) and (33). More precisely, if *a* > 0 (resp. *a* < 0) and *b* > 0, then at *β* = *β*\* there is a backward (resp. forward) bifurcation.

By evaluating the Jacobian matrix of system (17) at *E*_*dfe*_ and *β* = *β*\*, we get

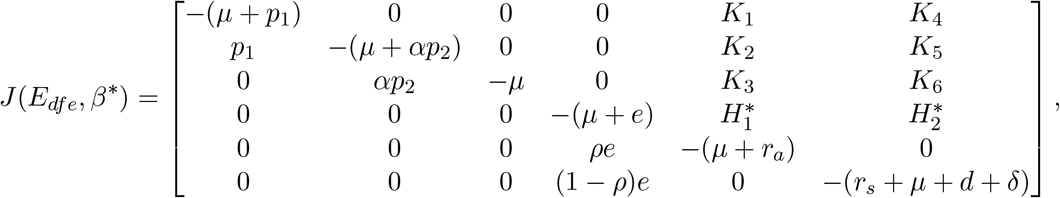

where

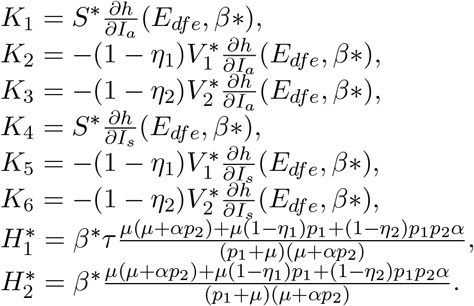

We observed that one of the eigenvalues of *J*(*E*_*dfe*_, *β*\*) is 0 and the remaining are negative. Hence, when *β* = *β*\* (when *R*_*v*_ = 1), the disease-free equilibrium is nonhyperbolic.

After some calculations we get:

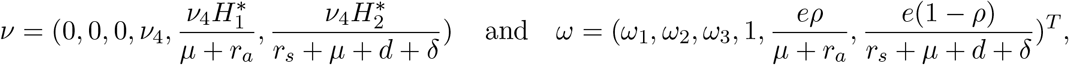

where

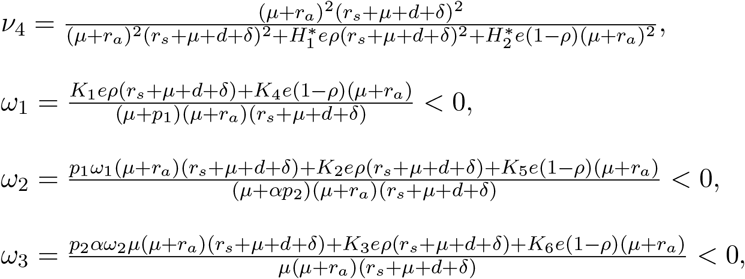

are a left and right eigenvector associated with the zero eigenvalue, respectively, such that *ν · ω* = 1. By considering only the nonzero components of the eigenvectors and computing the corresponding second derivatives of *f* we can explicitly compute the coefficients *a* and *b* as:

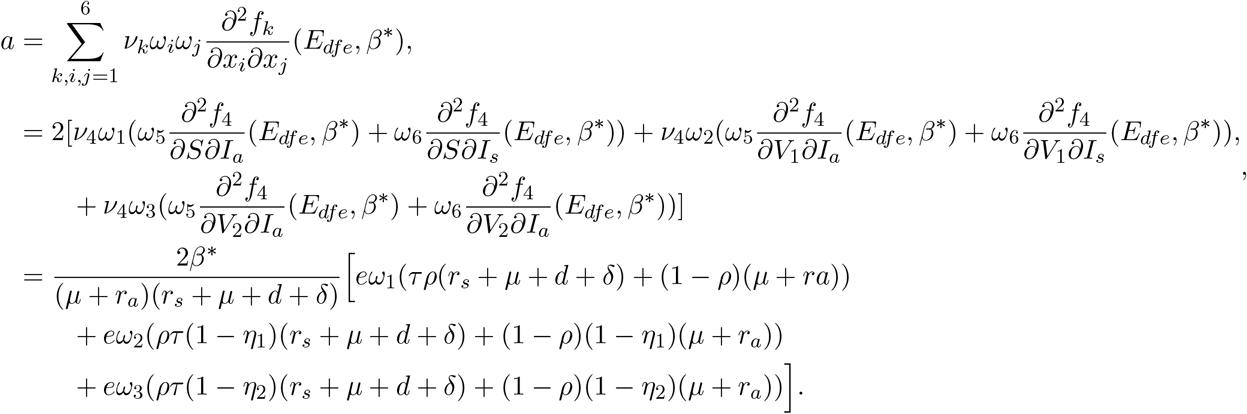

Since *ω*_1_, *ω*_2_ and *ω*_3_ are negative, it follows that *a* < 0 and

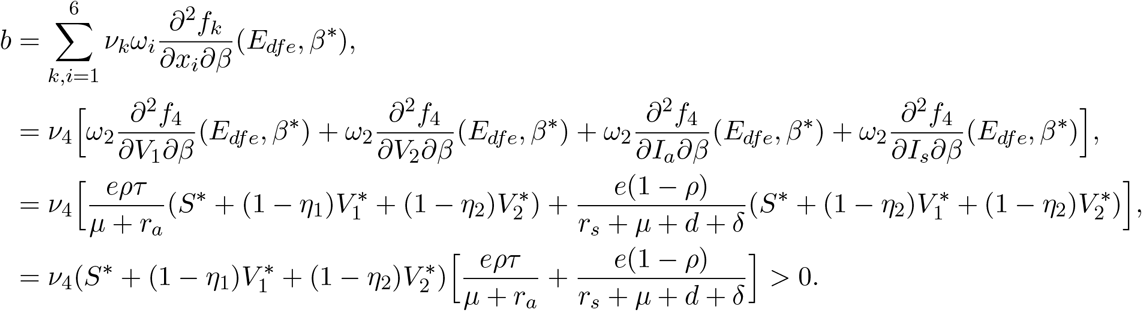

From the fact that *a* < 0 and *b* > 0, by the result of Castillo-Chavez and Song [7], as *R*_*v*_ passes through 1 a locally stable endemic equilibrium appears with the unstable disease free equilibrium. Therefore, model (17) exhibits a forward bifurcation at *R*_*v*_ = 1(see Figure 6). We summarize the above discussion with the following theorem.

#### Theorem 3.5.1

*The endemic equilibrium point, E*_*end*_ *of the model system* (17) *is locally asymptotically stable for R*_*v*_ > 1 *and the system exhibits a forward(or transcritical) bifurcation at R*_*v*_ = 1.

#### Remark 2

*From the bifurcation analysis and Theorem 3.4.1 for the full model (model with) we note that when R*_0_ = 1, *we have R*_*v*_ < 1 *in such case the disease free equilibrium is at least locally asymptotically stable*.

### 3.6 Sensitivity analysis

In what follows, we investigate the sensitivity analysis for the control reproduction number *R*_*v*_ to identify the parameters that have a high impact on disease expansion in the community. The sensitivity index with respect to a parameter *X*_*i*_ is given by a normalized forward sensitivity index [9],

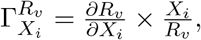

where, *X*_*i*_ represent the basic parameters. Hence,

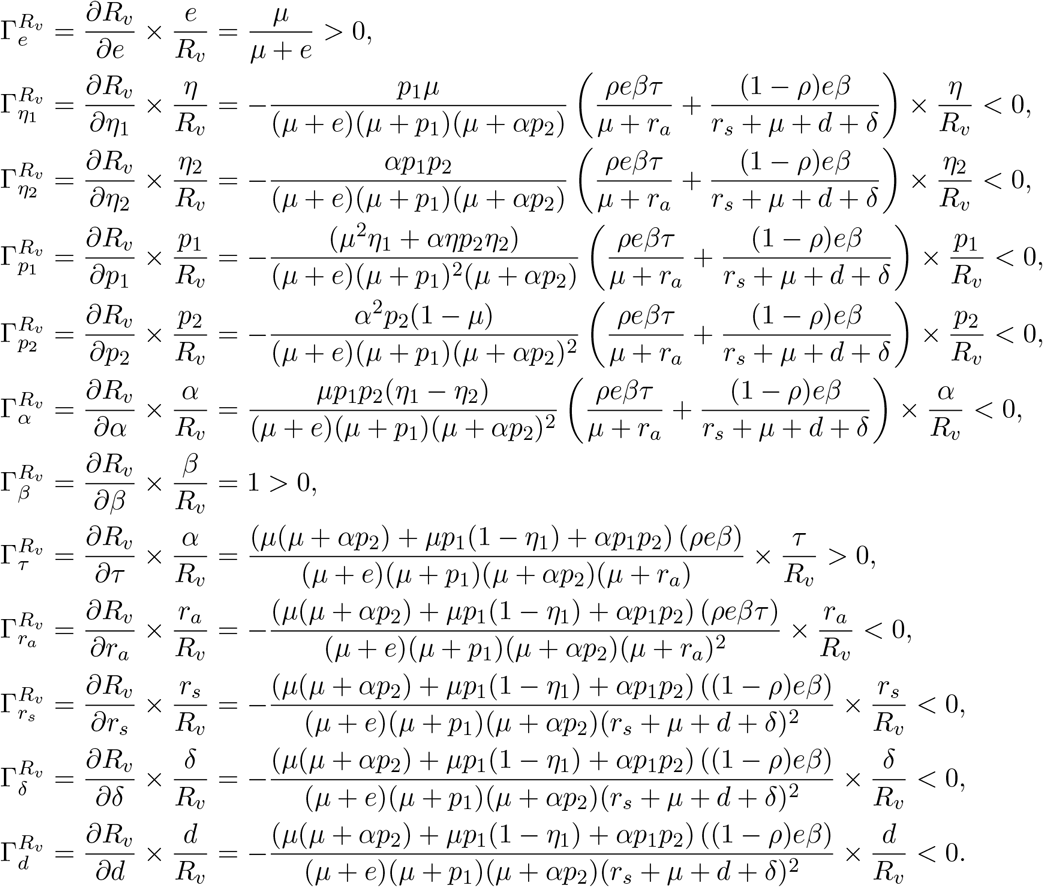

We summarize the sensitivity analysis indices of the reproduction number with respect to some parameters in Table 1.

**Table 1:** Sensitivity index table

| parameter | index |
| --- | --- |
| $e$ | +ve |
| $\beta$ | +ve |
| $\tau$ | +ve |
| $\eta_1$ | -ve |
| $\eta_2$ | -ve |
| $p_1$ | -ve |
| $p_2$ | -ve |
| $\alpha$ | -ve |
| $r_a$ | -ve |
| $r_s$ | -ve |
| $\delta$ | -ve |
| $d$ | -ve |

From Table1 the sensitivity indices with negative signs indicate that the value of *R*_*v*_ decreases when the parameter values are increased and the value of *R*_*v*_ increases when the parameter values are decreased, while sensitivity indices with positive signs indicate that the value of *R*_*v*_ increases when the parameter values are increased and the value of *R*_*v*_ decreases when the parameter values are decreased.

### 3.7 The role of vaccination

If there is no vaccination (i.e. *p*_1_ = *p*_2_ = 0), then *R*_*v*_ = *R*_0_. In such case disease elimination is possible if *R*_0_ < 1. and the disease will be endemic if *R*_0_ > 1. (Theorem 3.3.1). Suppose *R*_0_ > 1 and according to theorem 3.4.1, disease elimination is possible if *R*_*v*_ < 1. From

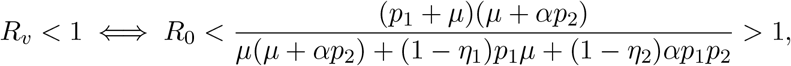

we get

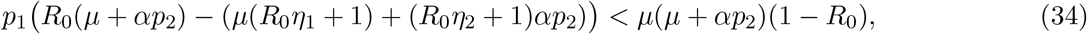

Since the right hand side of the inequality (34) is negative we must have

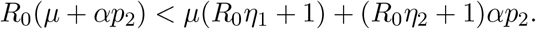

Therefore, *R*_*v*_ < 1 if and only if 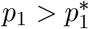. Where

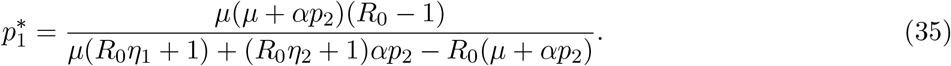

We call 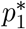 as a critical first dose vaccination rate.

Using the parameter values in the Table 2 the critical first dose vaccination can be calculated as 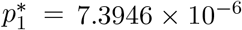. As it can be seen from the Figure 2, the control reproduction number will be less than one if 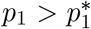. From epidemiological point of view to control the disease it is critical to increase the vaccination rate above 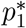.

**Table 2:** Parameter description and their baseline values used in the model (2).

| Parameter | Description | Value | Sources |
| --- | --- | --- | --- |
| $\pi$ | Recruitment rate | $4646 \text{ day}^{-1}$ | Calculated<br>Sec.4.1 |
| $\mu$ | Natural death rate | $\frac{1}{67.8 \times 365} \text{ day}^{-1}$ | Calculated<br>Sec.4.1 |
| $p_1$ | First dose Vaccination rate | $8.157 \times 10^{-7} \text{ day}^{-1}$ | Fitted |
| $p_2$ | Second dose Vaccination rate | $0.974 \text{ day}^{-1}$ | Fitted |
| $\beta$ | Transmission rate | $0.513 \text{ day}^{-1}$ | Fitted |
| $\tau$ | Infectivity factor for asymptomatic individuals | 0.116 | Fitted |
| $\eta_1$ | Efficacy of first dose vaccine | 0.8 | Fitted |
| $\eta_2$ | Efficacy of second dose vaccine | 0.95 | Fitted |
| $\alpha$ | Inverse of average time needed to take the second dose | $0.14 \text{ day}^{-1}$ | Fitted |
| $\rho$ | fraction of infections that become asymptomatic | 0.112 | Fitted |
| $e$ | Infection rate after incubation period | $0.2071 \text{ day}^{-1}$ | Fitted |
| $r_s$ | Recovery rate for individuals with symptom | $1.89 \times 10^{-7} \text{ day}^{-1}$ | Fitted |
| $r_a$ | Recovery rate for asymptomatic individuals | $0.0148 \text{ day}^{-1}$ | Fitted |
| $r_q$ | Recovery rate for quarantined individuals | $0.0356 \text{ day}^{-1}$ | Fitted |
| $r_h$ | Recovery rate for individuals in hospital | $0.213 \text{ day}^{-1}$ | Fitted |
| $\delta$ | Quarantine rate | $0.453 \text{ day}^{-1}$ | Fitted |
| $d$ | Disease induced death rate | $0.177 \text{ day}^{-1}$ | Fitted |
| $q_h$ | Hospitalization rate from quarantine | $0.999 \text{ day}^{-1}$ | Fitted |

**Figure 2:**
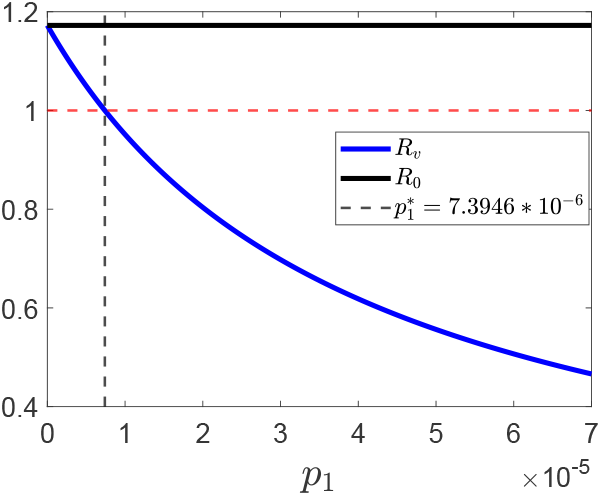
The role of the vaccination rate *p*_1_ on the control reproduction number *R*_*v*_ and the basic reproduction number *R*_0_. The red broken line is used to mark the horizontal line at 1. 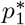 is calculated using the parameter values in the Table 2.

## 4. Numerical simulation and discussion

To justify the analytical results and explore additional important properties of the model, we fitted the model to real COVID-19 data of Ethiopia to fix the unknown parameters of the model and carried out numerical simulations using the MATLAB solver ODE45. In this section, we used the full model (2).

### 4.1 Parameter estimation

In this subsection, we will find the best values of unknown parameters in our model, with the so-called model fitting process. Here we shortly present how the fitting process works using the least square method. The system of equations (2) can generally be written as

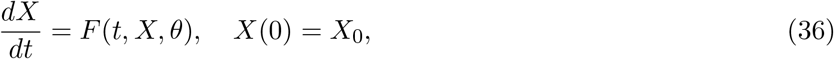

where *X* = (*x*_1_, *x*_2_, *· · ·, x*_*J*_) represents the state vector of the system with 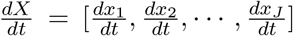, *J* is number of compartments in the population, *X*_0_ is a vector of initial values, *θ* = (*θ*_1_, *θ*_2_ *· · ·, θ*_*p*_) are unknown parameters of the system and *t* is the independent variable (time in our case) [19].

In order to estimate the unknown parameters *θ*, the state variable *X*(*t*) is observed at *N* time instants {*t*_1_, *t*_2_, *· · ·, t*_*N*_} so that we have

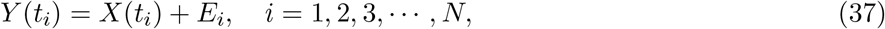

where *Y*(*t*_*i*_) is the observed values of the state variables at time instant *t*_*i*_ and 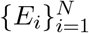 are the difference between the observed value *y*_*i*_ and the corresponding fitted value *x*_*i*_ (i.e. *E*_*i*_ = *y*_*i*_ − *x*_*i*_).The objective is to determine appropriate parameter values so that the sum of the squared errors between the outputs of the estimated model (*X*(*t*)) and the observed data (*Y*(*t*)) are minimized.

The best fit was achieved by searching for the set of parameters objective function 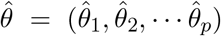 which satisfies the objective function

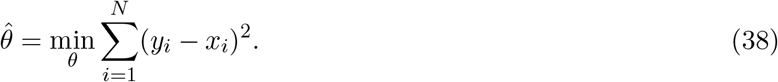

To find the best fit parameters for our model which satisfies the equation (38), we used the nonlinear curve fitting method with the help of ‘lsqcurvefit’, MATLAB built in function. Lsqcurvefit is an optimization toolbox which solves nonlinear data-fitting problems in the least-squares sense. In our case the number of parameters, *p*, to be estimated is 16. We fitted our model to the real data of COVID-19 daily cumulative confirmed cases and vaccinated population of Ethiopia from May 01, 2021 to January 31, 2022, which is available online by Our World in Data [23]. Two of the parameter values are estimated from literature: according to the data by Worldometer, the Ethiopian average life expectancy at birth for the year 2021 and the approximate total population is 67.8 and 114963588 respectively [34]. Therefore, the natural death rate of individuals per day is calculated as the reciprocal of the life expectancy at birth times days in a year, given by 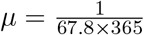. We approximated the recruitment rate from 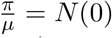 (Initial population). Hence we found *π* = *μ* × *N* (0) = 4646 individuals per day [18, 17]. In the estimation process of the rest parameters the following initial conditions are used: from the data in Our World in Data we have *I*_*s*_(0) = 620, *V*_1_(0) = 20385, *R*(0) = 946 and *D*(0) = 21. Where *t* = 0 corresponds to May 01, 2021. We assumed 80% of COVID-19 infected individuals become asymptomatic. Therefore we estimated *I*_*a*_(0) = 620*/*0.8 = 775. We also assumed *E*(0) = 1400, which is approximately equal to the sum of the symptomatic and asymptomatic cases, and *V*_1_(0) = *Q*(0) = *H*(0) = 0. Hence, the initial susceptible population is taken as *S*(0) = *N*(0) − (*V*_1_(0) + *V*_2_(0) + *E*(0) + *I*_*a*_(0) + *I*_*s*_(0) + *Q*(0) + *H*(0) + *R*(0)).

The best fit to the daily cumulative COVID-19 confirmed cases and vaccination through our model is shown in Figure 3 and it can be observed that the estimated parameters for the cumulative daily cases is well fitted as compared the observed data. The estimated and calculated parameter values are given in Table 2. Using these parameters, we calculated *R*_0_ = 1.17 and *R*_*v*_ = 1.15. The estimated value of the basic reproduction number is greater than 1 which is similar as the study for Ethiopia in [17] in which they estimated *R*_0_ = 1.0029. In the same study the estimated transmission rate is *β* = 0.88 which is greater than our case, which is can be expected due to in our case we have vaccination as a control strategy. Thus, Apart from the uncertainty in the parameter values due to the model’s complexity, the estimated parameters can represent the situation in Ethiopia at the time the data is collected.

**Figure 3:**
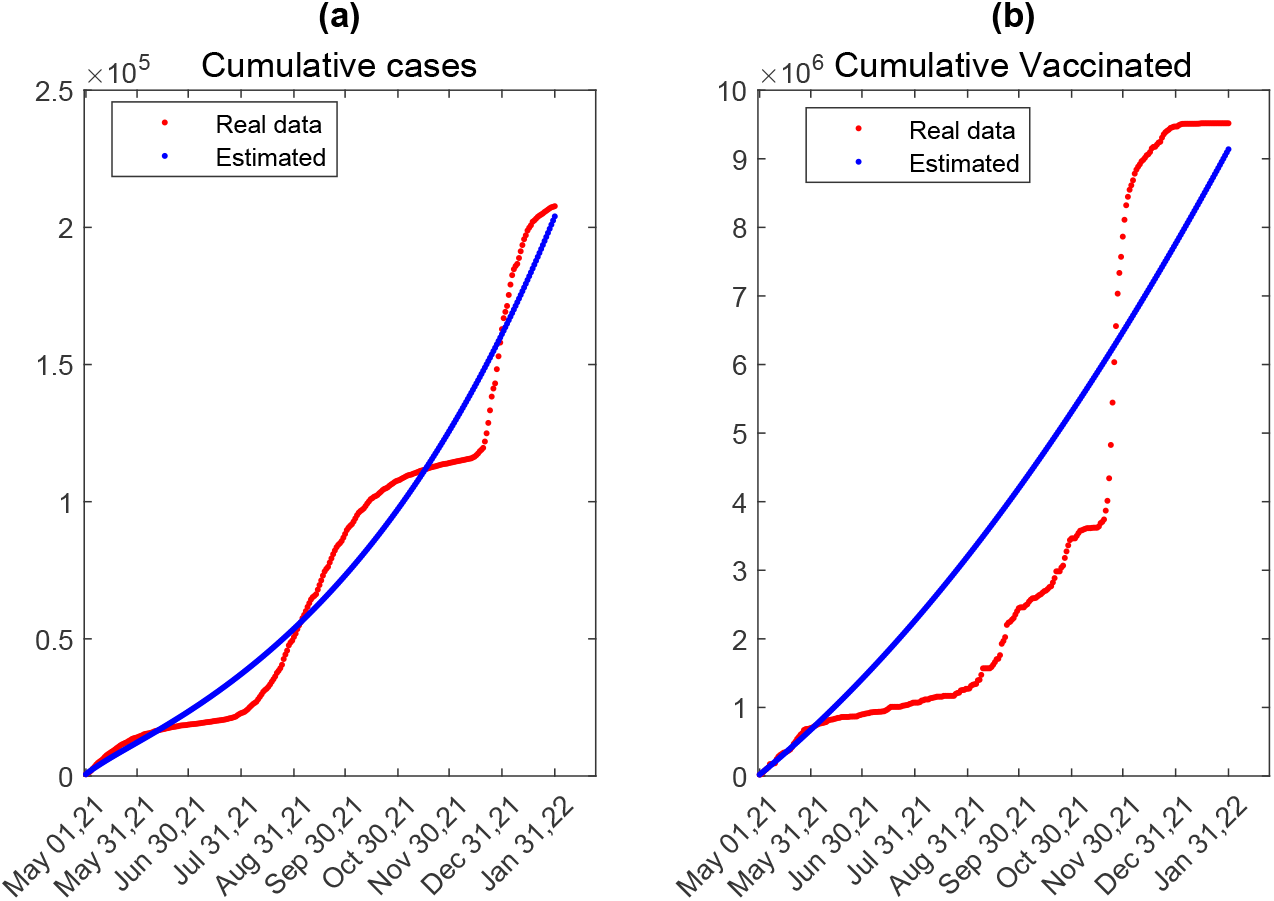
The fitted data to the reported cumulative cases (panel (a)) and cumulative vaccinated(panel (b)) using the model (2) for Ethiopia from May 01, 2021 to January 31, 2022.

### 4.2 Long-term dynamics of the model

Figure 4, panels (*a*) and (*b*) (for time interval [9000, 30000]) shows the local stability of the endemic equilibrium *E*_*end*_ = [3.77 × 10^−7^, 225, 6.91 × 10^5^, 1.49 × 10^4^, 2.334 × 10^4^, 4.36 × 10^3^, 1.632 × 10^3^, 4.181 × 10^3^, 3.201 × 10^7^] for *R*_*v*_ = 2.98 > 1. Panels (*c*) and (*d*) portrays the stability of the disease free equilibrium,*E*_*dfe*_ = [1.127 × 10^8^, 673.9, 2.2741 × 10^6^, 0, 0, 0, 0, 0, 0], for *R*_*v*_ = 0.556 < 1. These results support our analytical results in section 3 of Theorem 3.4.2 and 3.5.1. For better use of spacing and view we didn’t include the plot for *E* compartment, but the dynamics of this state variable converges to its equilibrium point. The convergence to the endemic equilibrium is through damped oscillation, which shows the disease may re-emerge. Such long-term oscillatory dynamics are consistent with the findings of an Indian study [16], suggesting that COVID-19 could become a seasonal disease.

**Figure 4:**
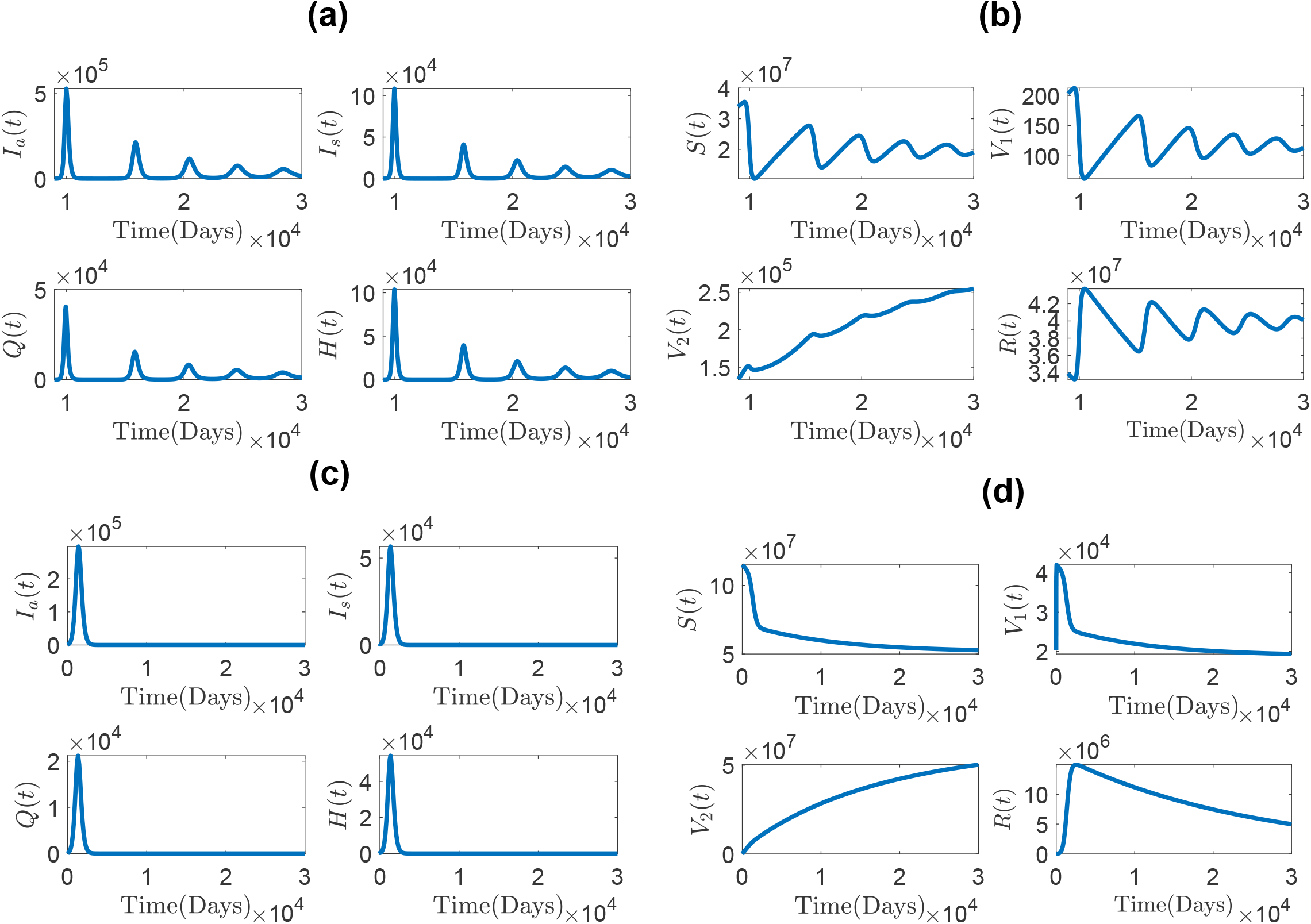
Local stability of the endemic equilibrium for *R*_*v*_ = 2.98 > 1(infected compartments, panels (*a*), and non infected compartments, panel (*b*)) and local stability of the disease free equilibrium for *R*_*v*_ = 0.556 < 1 (infected compartments, panel (*c*), and non infected compartments, panel (*d*).) *τ*_1_ = 0.6 and *p*_1_ = 5 × 10^−5^ is used for panels (*a*)&(*b*) and (*c*)&(*d*) respectively and other parameter values are given in Table 2.

When *R*_*v*_ = 1 an exchange of stability (forward bifurcation) arises, This property is shown in Figure 6. Which shows the disease persists in the population if the reproduction parameter excedes the threshold value.

### 4.3 Variation of *R*_*v*_ and *R*_0_ with respect to some important parameters

An important parameter in modeling infectious disease transmission is the reproduction parameter which measures the potential spread of an infectious disease in a community, in our case we have a control reproduction parameter, *R*_*v*_. In particular, if *R*_*v*_ < 1 the disease dies out and if *R*_*v*_ > 1 the disease persists in the population. Therefore, reducing such parameter below the critical value *R*_*v*_ = 1 is important. In our model, reducing the transmission rate *β* and infectivity factor of asymptomatic individuals, *τ* will help reduce *R*_*v*_ from unity, Figure 5 panels (*a*) and (*b*). It is worth noting that the influence of the second dose vaccination rate on varying the control reproduction number is minimal. Keeping parameters other than the transmission rate *β* constant as in the Table 2, *R*_*v*_ < 1 if *β* < 0.4464 (See Figure 5 panel (*a*) or (*c*)). If *τ* < 0.0764, then *R*_*v*_ < 1 fixing other parameters constant, Figure 5 panel (*b*).

**Figure 5:**
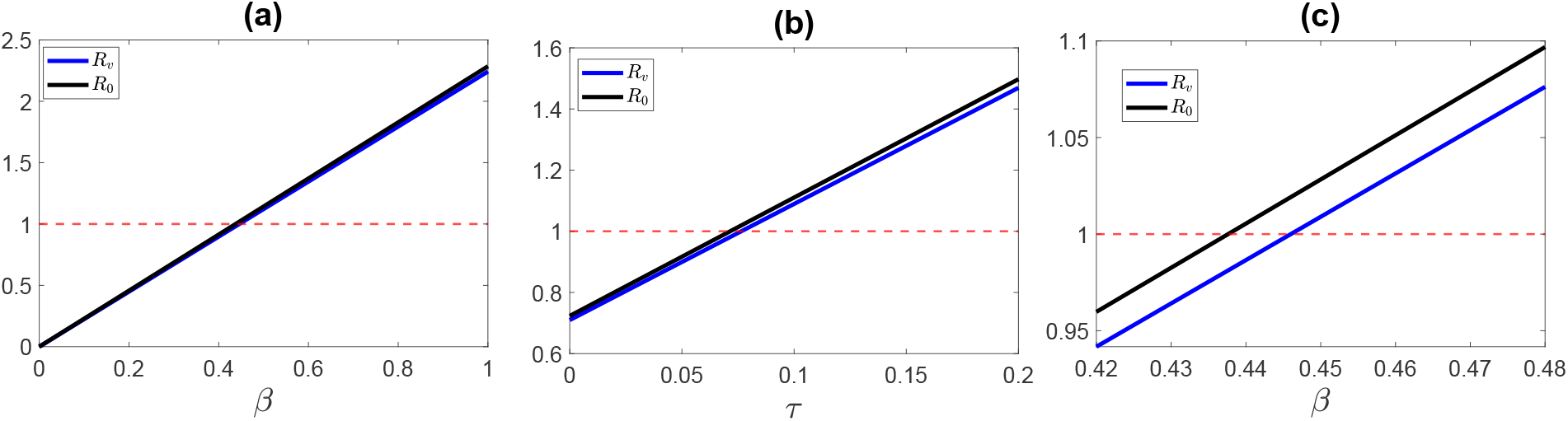
Variation of *R*_*v*_ with respect to : the transmission rate *β*, panel (*a*) and infectivity factor of asymptomatic individuals *τ*, panel (*b*). Panel (*c*) shows the zoom plot of panel (*a*). The red doted line is used to mark the line at one. Other parameter values are given in Table 2.

**Figure 6:**
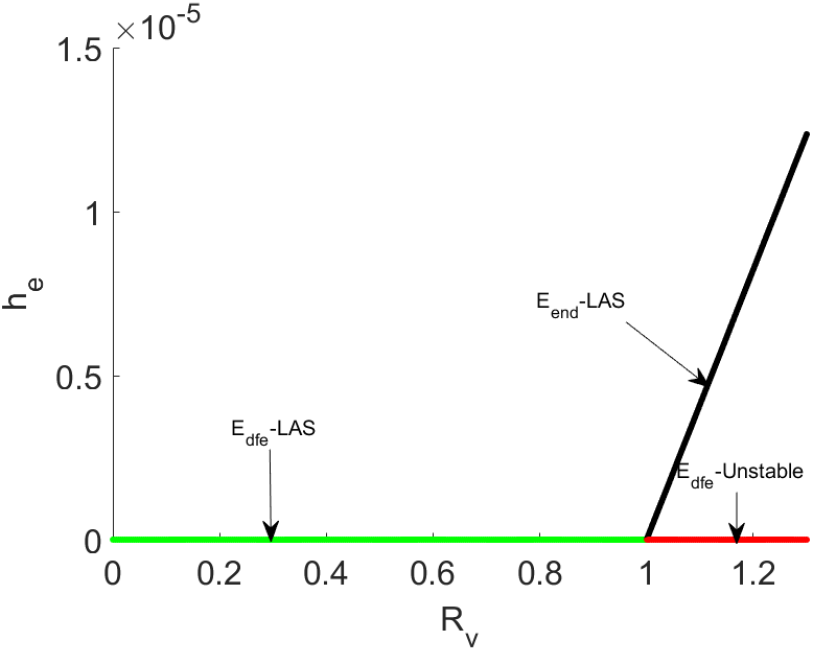
Transcritical bifurcation of model (2) when *R*_*v*_ = 1.

### 4.4 The impact of transmission rate

In this and subsequent subsections, we say infectious population to refer to the sum of the population in symptomatic and asymptomatic classes per time (*I*_*a*_(*t*) + *I*_*s*_(*t*)). This is due to the fact that in our model we assumed people in these two compartments are potential transmitters of the disease. Unless explicitly mentioned, when we say vaccinated individuals, it refers to the total number of individuals vaccinated either with the first dose or the second dose per unit time (*V*_1_(*t*) + *V*_2_(*t*)). Figure 7 shows the role of the transmission rate *β* on the dynamics of the infectious, vaccinated, and hospitalized classes. A decrease in the transmission rate results in a prevalence decrease. When the transmission rate is equal to 0.55 *days*^−1^ the prevalence reaches a high peak of 1424101, but by decreasing it to *β* = 0.49 *days*^−1^ (below the fitted value) the infectious peak can be decreased to 410094 Figure 7 panel (*a*). This shows that if we can further decrease the transmission rate, it is possible to achieve an infectious number of insignificant value and eradication of the disease. When the transmission rate is small, a small number of people will be infected, which means the number of people in the susceptible class will be large, hence the number of vaccinated people will rise, Figure 7, panel(*b*). The burden of hospitalization can be decreased by decreasing the transmission rate. As it can be seen in Figure 7, panel(*c*), when the infectious population is high, correspondingly we have a large number of individuals in the hospital and vice versa.

**Figure 7:**
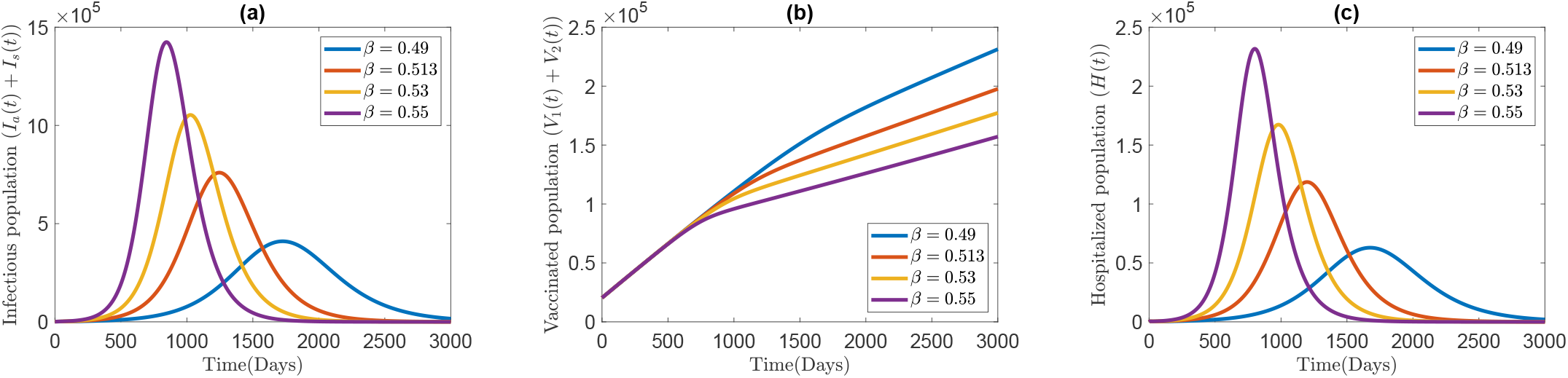
The effect of transmission rate *β*. Panel (a): infectious population *I*_*a*_(*t*) + *I*_*s*_(*t*), panel (b): Vaccinated population, *V*_1_(*t*) + *V*_2_(*t*), and panel (c) hospitalized individuals. Other parameter values are given in the Table 2.

### 4.5 The impact of first dose vaccination rate

Figure 8 shows the role of the first dose vaccination rate on the dynamics of infectious, vaccinated and hospitalized population. Increasing this vaccination rate results in a decrease of infectious and hospitalized population Figure 8 panels (*a*)&(*c*). For example when *p*_1_ = 8.16 × 10^−7^ *day*^−1^ the infectious population reaches a high peak of value 759544 and hospitalized peak of 118624 individuals. If we are able to increase the rate to *p*_1_ = 8.16 × 10^−5^ *day*^−1^ the above peaks will decrease to 171226 and 26151 of infectious and hospitalized individuals respectively. Such a decrease in prevalence is achieved with high proportion of vaccinated individuals in the population Figure 8 panel (*b*). Simulation results shows that the role of the second dose vaccination rate, *p*_2_ and time delay between the two doses, *α* doesn’t have significant impact on the dynamics. From the formulation of the model, every one who got the first dose and not infected is assumed to get the second dose and therefore will be transferred to *V*_2_ class after an average time of 1*/α* hence the role of the vaccination is visible when *p*_1_ varies. If health officials attempt to encourage people to get the second dose of the vaccination, the prevalence will drop dramatically.

**Figure 8:**
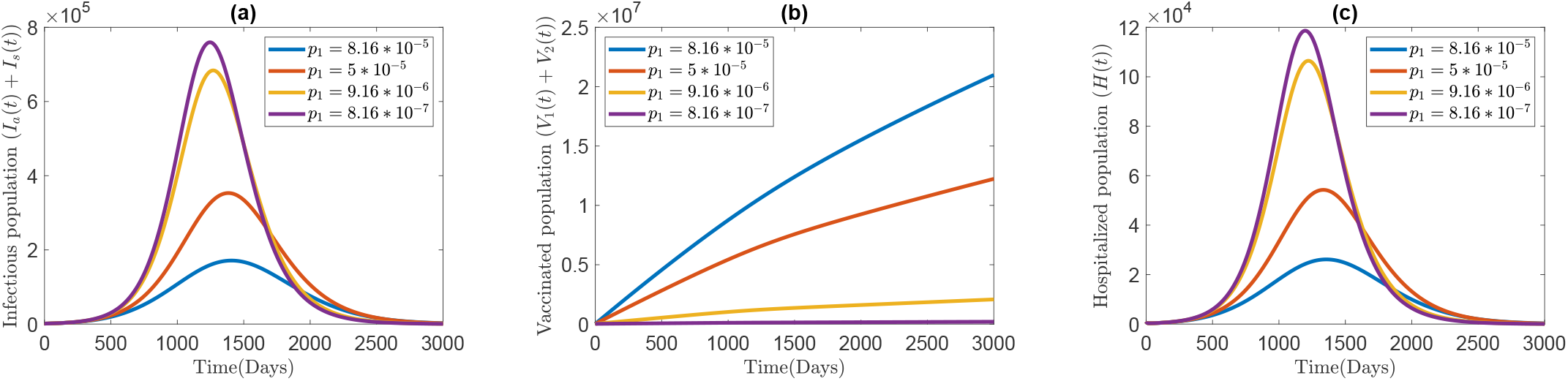
The impact of the first dose vaccination rate *p*_1_: on the dynamics of infectious population,panel (*a*), vaccinated population, panel (*b*), and hospitalized population, panel (*c*). Other parameter values are given in the Table 2.

### 4.6 The impact of the infectivity factor of asymptomatic individuals

According to the study [28], asymptomatic cases of COVID-19 are a potential source of substantial spread of the disease within the community and one of the results found was people with asymptomatic COVID-19 are infectious but might be less infectious than symptomatic cases. Since the majority of COVID-19 infected individuals become asymptomatic, even if they are less infectious than the symptomatic individuals, their role in spreading the disease might be significant. Figure 9 proves this hypothesis. As the infectivity factor increases, we observed a rise of the infectious population to a relatively high pick (2799983 infectious for *τ* = 0.2) Figure 9, panel (*a*), which is not observed in the impact of other parameters, like *β*. Decreasing the infectivity factor decreases the infectious population significantly. As observed in other plots here also the increase of infectious population will result in increase in the number of hospitalized individuals and vice versa Figure 9 panel (*c*). The increase in the infectivity factor *τ* makes more people to be infected from vaccinated compartments which results in a decrease in the number of vaccinated individuals, Figure 9 panel (*b*). Therefore the number of vaccinated individuals is inversely proportional to the infectivity factor. Detection of Asymptomatic individuals (for example: by contact tracing) and isolating them may reduce their infectiousness.

**Figure 9:**
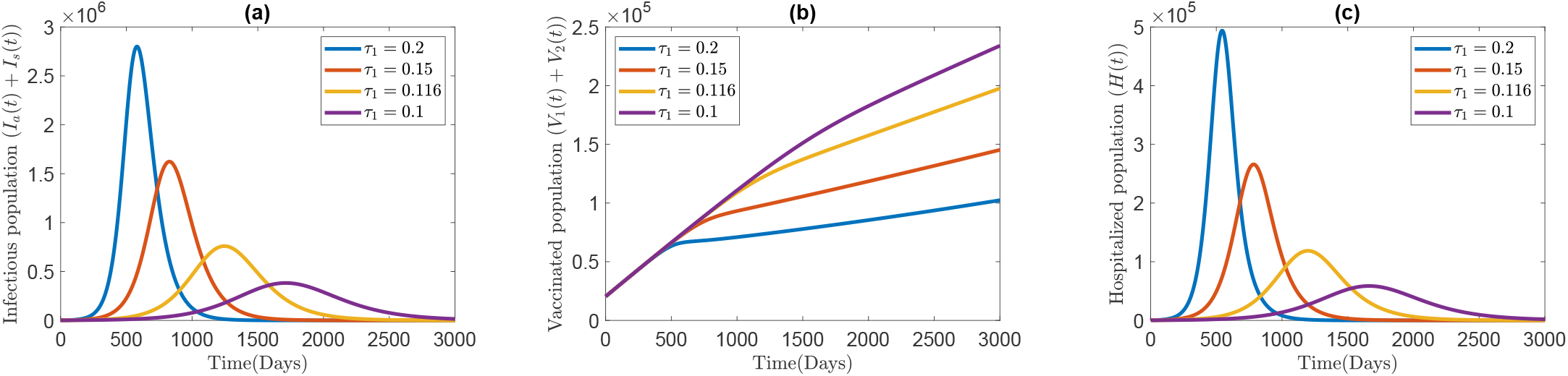
The impact of the infectivity coefficient of asymptomatic population, *τ*_1_ on the dynamics of infectious population,panel (*a*), total vaccinated population, panel (*b*), and hospitalized population, panel (*c*). Other parameter values are as in the Table (2).

## 5. Prediction of cumulative vaccine dose administered with respect to the first dose vaccination rate

Most of COVID-19 vaccines approved by WHO are being offered in two doses and a booster. In Ethiopia Sinopharm, AstraZeneca, Johnson and Johnson/Janssen, and Pfizer-BioNTech vaccines are being used. From these vaccines except Johnson&Johnson/Janssen all are being given in two doses. The total number of COVID-19 vaccine dose administered from May 01, 2021 to January 31, 222 (276 days) is 9517539. Using the fitted parameters, our model estimates this number by 9152542 vaccine doses (See, the highlighted row third column of Table 3). If the first dose vaccine administration rate remains the same for the next two years, (*i*.*e* after 1006 days) 66483093 number of vaccine doses will be administered. According to World Population Review projection, Ethiopian population in 2024 will be 126.8 million [33]. Since a person can get vaccinated with two doses, we can approximate the number of people vaccinated with at least one dose by 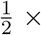 number of vaccine dose administered. This means 33241546 number of people (Approximately 26% of the total population (in 2024)) will get at least one dose of COVID-19 vaccination. Increasing *p*_1_ to 3.16 × 10^−6^ *days*^−1^ it can be achieved, after two years, 199688874 number of administered vaccine doses. Which is equivalent to 99844437 number of people (approximately 79% of the total population in the year 2024) can get at least first dose (see fourth row of Table 3). It needs a lot to work on increasing the vaccination rate beyond the critical value 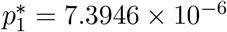 so that *R*_*v*_ < 1.

**Table 3:** Values of: Control reproduction number (second column), cumulative vaccine administered at the end of the parameter fitting time (third column) and Predicted number of cumulative vaccine to be administered (fourth column). For different values of *p*_1_. Other parameter values are given in Table 2. The light Cyan shaded row is for the base line *p*_1_ value.

| $p_1$ | $R_v$ | Vaccine dose administered in $[0, 276]$ days (Interval of fitting time) | Predicted after two years ($[0, 1006]$ days interval) |
| --- | --- | --- | --- |
| $8.157 \times 10^{-7} \text{ day}^{-1}$ | 1.15 | 9152542 | 66483093 |
| $9.16 \times 10^{-7} \text{ day}^{-1}$ | 1.147 | 9588497 | 72169187 |
| $1.16 \times 10^{-6} \text{ day}^{-1}$ | 1.141 | 10652193 | 86042042 |
| $3.16 \times 10^{-6} \text{ day}^{-1}$ | 1.09 | 19369216 | 199688874 |

## 6. Conclusion

In this study, we used a compartmental model for COVID-19 transmission with vaccination. We divided the vaccinated portion of the population into two: Vaccinated with the first dose and fully vaccinated (those who got the two doses). Using the next generation matrix, we found a reproduction number which exists when vaccination is in place. We called this parameter the control reproduction number and denoted it by *R*_*v*_. We calculated the disease-free and endemic equilibrium of model (2) and showed that the disease-free equilibrium *E*_*dfe*_ is globally asymptotically stable if the control reproduction number *R*_*v*_ < 1 and unstable if *R*_*v*_ > 1. We performed a center manifold analysis based on the method mentioned in Castillo-Chavez and Song[7] and found that the model exhibits a forward bifurcation at *R*_*v*_ = 1, which ensures the nonexistence of the endemic equilibrium below the critical value, *R*_*v*_ = 1 and the unique endemic equilibrium which exists for *R*_*v*_ > 1 is locally asymptotically stable. From epidemiological point of view this implies that the disease dies out if the control reproduction number is below the threshold quantity and it persists in the population if greater. This informs public health policy makers to work on reducing the control reproduction number so as to make it less than unity. We performed a sensitivity analysis from which we observed that the model is sensitive to *p*_1_, *p*_2_, *δ* with negative sign and *β, τ* with positive sign. This shows that increasing the vaccination and quarantine rate and decreasing the transmission rate and infectivity factor of asymptomatic individuals will reduce the disease burden.

We performed model fitting to the Ethiopian real COVID-19 data for the period from May 1, 2021 to January 31, 2022 to estimate the unknown parameters in the model. In the numerical simulation section, we validate our analytical analysis regarding the stability of the disease-free and endemic equilibrium with respect to the parameter *R*_*v*_. We also examined the role of some important parameters on the dynamics of the disease and arrived at the following points: Reducing the transmission rate and the infectivity factor of asymptomatic individuals will greatly help in reducing the infection burden. Increasing the first dose vaccination rate has a high impact in reducing the infection. Furthermore, simulation results show that the second dose vaccination rate has no significant effect on the dynamics of the infectious population.

In addition to this, we also predicted the cumulative vaccine dose administered by changing the first dose vaccination rate. In this prediction, if we increase *p*_1_ to a value 3.16 × 10^−7^ *day*^−1^ after two years, the total vaccine dose administered will reach 1996888974, which will cover approximately 79% of the total population. Therefore, from the numerical simulation and analytical analysis, we summarize that it will be essential to reduce the transmission rate, infectivity factor of asymptomatic cases and increase the vaccination rate beyond the critical value 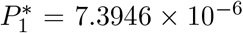, quarantine rate to control the disease. As a future work, we will point out that this model can be extended by including additional interventions (for example, non-pharmaceutical interventions), by considering the behavioural aspect and via optimal control problem. We would also want to point out that the model can be studied further using fractional order derivatives and the findings obtained can be compared.

**This article has been accepted for publication in the Discrete Dynamics in Nature and Society journal**.

## Data Availability

All data produced in the present study are available upon reasonable request to the authors.

https://ourworldindata.org/

## Data Availability

Data will be available on request.

